# Spine-Related Health Care Utilization and Costs Following Orthobiologic Injection Versus Lumbar Surgery for Degenerative Spine Conditions

**DOI:** 10.64898/2026.03.31.26349877

**Authors:** Trevor A. Lentz, Joshua Burrows, Amanda Brucker, A. Ian Wong, Laura Qualls, Rekha Divakaran, Chris Centeno, Tim Suther, Laine Thomas

## Abstract

**Background:** Lumbar fusion and decompression procedures are widely used for degenerative spine conditions but are associated with substantial health care costs and variable outcomes. Orthobiologic treatments, including platelet rich plasma (PRP) and bone marrow aspirate concentrate (BMAC), have emerged as less invasive options for select patients who meet surgical criteria. However, concerns remain that orthobiologic care may delay rather than avert surgery, potentially increasing downstream utilization and costs. Comparative evidence on real world utilization and costs is limited.

**Methods:** We conducted a retrospective, observational study using linked commercial insurance claims and a national orthobiologic treatment registry. Adults with lumbar degenerative disc disease (DDD) who met criteria for lumbar fusion or laminectomy, foraminotomy, discectomy, and facetectomy (LFDF) procedures, and who received PRP injection (with or without BMAC) or surgery between 2016 and 2023 were included. Two comparisons were evaluated: PRP versus lumbar fusion and PRP versus lumbar decompression procedures. Propensity score matching was used to balance cohorts on demographic characteristics, comorbidities, spine related diagnoses, prior health care use, and severity proxies. Outcomes included spine-related health care resource use and aggregate costs at 12 and 24 months, with exploratory analyses at 36 and 48 months. Costs were estimated using multiple approaches, including Medicare based estimates and commercial payer methods.

**Results:** After matching, 133 patients receiving PRP were compared with 2,560 patients undergoing fusion, and 198 patients receiving PRP were compared with 3,960 patients undergoing LFDF. Rates of subsequent spine surgery following PRP were low and below cell suppression thresholds through 24 months, with similar findings in exploratory longer-term analyses. Compared with surgical cohorts, patients receiving PRP had lower rates of postoperative imaging, home health services, and outpatient visits, with no consistent differences in opioid use, magnetic resonance imaging, or physical therapy. At 12 and 24 months, mean aggregate costs were significantly higher for fusion and LFDF cohorts across most costing methods. Cost differences were largest for fusion comparisons and were driven primarily by index procedure costs and higher reoperation and imaging rates in surgical cohorts. Findings were generally consistent across sensitivity and exploratory analyses.

**Conclusions:** Among select patients with degenerative spine conditions who meet surgical criteria, PRP was associated with lower health care utilization and substantially lower costs compared with lumbar fusion or LFDF, without evidence of increased progression to surgery. These findings support consideration of orthobiologic options for appropriately selected patients when surgery is not the only viable treatment option. Limitations include selection bias, absence of patient reported outcomes, and claims-based severity measures.

## BACKGROUND

Lumbar fusion and decompression procedures including laminectomy, foraminotomy, discectomy, and facetectomy (collectively referred to as LFDF in this paper) are among the most frequently performed surgeries for degenerative spine conditions and are a major driver of health care expenditures in the United States.^9^ These interventions are intended to alleviate pain and restore function in patients with spinal instability, stenosis, or disc pathology. However, up to 25% of patients continue to experience chronic postsurgical pain requiring opioid use, with reoperation rates ranging from 6-27%.^14,24,29,30^ Decompression procedures for degenerative spine disease have demonstrated mixed results, with some studies showing limited advantage over nonoperative care.^2,33^ Surgery remains necessary for severe cases, especially with neurological deficits or structural deformity. But their widespread use for degenerative conditions has raised concerns about appropriateness and value, particularly when less invasive options have not been fully explored or when the expected benefit of a procedure is uncertain.^27^

As health systems and payers seek to lower costs and improve outcomes for spine care, interest in less invasive, cost-conscious alternatives has grown. Orthobiologic therapies represent one such option that carry fewer inherent risks (e.g., infection, anesthesia complications, nerve injury) and shorter recovery times than surgery.^3,7,11,17,18,23^ Platelet-rich plasma (PRP) and bone marrow aspirate concentrate (BMAC) have emerged as biologic interventions with potential to modulate inflammation and promote tissue healing.^10,20^ PRP delivers concentrated platelets that release growth factors and cytokines, while BMAC provides mesenchymal stem cells (MSCs), hematopoietic cells, and anti-inflammatory mediators that may support disc and bone repair.^15^ MSCs exhibit multipotent properties that enable differentiation into musculoskeletal tissues, offering theoretical benefits for degenerative spine conditions.^5,19,35^ Intradiscal PRP and bone marrow concentrate have shown statistically significant improvements in pain and function up to two years following treatment for discogenic low back pain.^25,26,31,32^

The adoption of PRP and BMAC remains low, in part because these treatments are not often covered by insurance and very few studies have directly compared outcomes between these treatments and common alternatives like surgery. Evidence suggests these therapies may improve pain and function in patients with lumbar degenerative pathology^25,26,31,32^, yet their long-term impact on disease progression and health care utilization is unclear. Orthobiologic procedures are often promoted as alternatives to surgery for some patients, but concerns remain that they may only postpone inevitable operations while increasing costs through repeat injections and adjunctive services (e.g., physical therapy, chiropractic care), raising questions about long-term value. As payer policies evolve and spine care costs escalate, understanding how orthobiologics compare to surgery in terms of health care use and cost is critical for informed decision-making.

This study examines spine-related utilization and costs among patients with lumbar degenerative conditions who meet surgical criteria for fusion or LFDF and select orthobiologic injection (PRP with or without BMAC) or receive surgery. The primary endpoint is aggregate cost over 12 and 24 months with exploratory analyses at 36 and 48 months post procedure. By leveraging real-world data, we aim to determine whether orthobiologics function as a cost-conscious alternative to surgery or contribute to increased health care burden.

## METHODS

### Study population

This is a retrospective, observational study using health care claims data to compare cost and resource utilization outcomes for patients undergoing 1) autologous PRP with or without BMAC (hereafter, PRP) treatment versus spinal fusion, and 2) autologous PRP versus LFDF. All procedures occurred between 2016 and 2023. All patients were enrolled in a commercial health insurance plan.

### Data sources

Health care use data including facility, provider, and pharmacy claims and financial fields for all associated claims data was obtained from a nationwide commercial insurance provider, through their research data platform. Details on PRP procedures were extracted from the Regenexx patient registry and included procedure date, treated joint, treatment location, and injection type. Aside from these procedure-specific variables, all covariates and outcome measures were sourced from the commercial claims database for all comparator groups. Procedural data from Regenexx were linked to the claims dataset using a privacy-preserving, token-based method performed by Datavant, Inc. No identifiable information was exchanged between organizations other than service dates, and the final analytic dataset was de-identified by expert determination. Analyses were conducted within a secure, cloud-based enclave designed to prevent data exfiltration; only privacy-screened summary outputs were allowed to leave the environment. This study was reviewed and approved by the Duke IRB (Pro00116431).

### Eligibility criteria

Patients were eligible if they 1) received an eligible orthobiologic treatment (PRP; Regenexx, Des Moines, IA) or underwent surgery (Supplementary Appendix 1); 2) had a procedure between 2016 and 2023; 3) were age 18 years or older at the time of the procedure; 4) had commercial coverage from 12 months prior to procedure to at least 12 months after procedure; 5) had at least one outpatient encounter (claim) in the 12 months prior to the procedure and at least one outpatient encounter (claim) in the 12 months after the procedure; and 6) had an elective procedure that was preceded by a clinical diagnosis that is consistent with degenerative disc disease (DDD) of the lumbar spine (lumbar disc degeneration or spondylosis, spondylolisthesis, stenosis, myelopathy, or radiculopathy). Additional details are in Supplementary Appendix 1.

### Conditions of the lumbar spine compatible with DDD

Patients were excluded if they had 1) multiple birthdates or sexes in the commercial payer data; 2) specific ineligible medical conditions (Supplementary Appendix 1) within the 12 months prior to the procedure, e.g., inflammatory syndromes, traumatic injuries, bone cancer; 3) previous spinal surgery or radiofrequency ablation during the same period; 4) musculoskeletal complications or related disorders in the prior year; or 5) evidence of receiving a monoclonal antibody drug within three months before the procedure. For the PRP cohort, we excluded patients with evidence of corticosteroids (oral or injection) within 2 months prior to the procedure as this is contra-indicated for use with PRP. If there were multiple eligible procedures for a patient, we took the first that met the eligibility criteria.

### Cohorts and comparators

The comparators of interest were: (1) spinal fusion versus PRP in patients who would have been a good candidate for both spinal fusion and PRP, and (2) LFDF versus PRP in patients who would have been a good candidate for both LFDF and PRP. Details of PRP and PRP treatments, including their preparation and administration, are provided elsewhere.^6,13^ In practice, whether a patient is a good candidate for these procedures is based on objective criteria as well as subjective judgement.

A base level of comparability was established by the inclusion criteria that requires evidence of lumbar DDD (see eligibility criteria). Conditions compatible with lumbar DDD were: disc degeneration or spondylosis, spondylolisthesis, stenosis, myelopathy, radiculopathy. Conditions not compatible with DDD if occurring in isolation (not with conditions above): herniated disc/displacement, disc disorders, low back pain (these would tend to not be managed surgically or with PRP). To be included in the lumbar fusion surgical cohort, patients had to meet study eligibility criteria and undergo lumbar fusion, which was determined using CPT codes. To be included in the LFDF surgical cohort, patients had to meet study eligibility criteria and undergo a LFDF procedure.

Second, we used propensity score matching to refine comparability. Some orthobiologic procedures may be compatible with multiple surgical options. Which one would be most likely may be definitive for some patients, but others may be compatible with multiple surgical alternatives. Therefore, we will use propensity score matching to identify patients receiving PRP who are compatible with spinal fusion and then separately identify patients receiving PRP who are compatible with LFDF.

### Confounders

We pre-specified a list of potential confounders thought to directly impact outcomes or potentially be a surrogate for direct causal factors. For derived variables that involve diagnosis or procedures codes, we included all such in the 12-month period prior to the date of the index procedure (excluding the date of the index procedure itself). Demographic information included age, sex, and geographic region. Health related characteristics included spine conditions, other musculoskeletal conditions not exclusive to the spine, comorbidities, pain medication prescriptions, intervention history, health seeking behavior, and other severity proxies (e.g., number of outpatient encounters due to spine condition). Variables were selected for propensity score matching prior to seeing any analysis of outcome data. That is, we prioritized variables that were different between the treatment groups and had a plausible impact on post-procedural cost outcomes (list in Supplementary Appendix 1).

Variables that were rare, well-balanced, or highly collinear with included covariates were not directly included within propensity score matching to preserve precision, given the finite sample size. We report balance diagnostics (standardized mean differences) on all candidate confounders, regardless of whether they were included within propensity score matching. A few clinically relevant variables were not treated as confounders because they were closely linked to surgical choice. For example, diagnostic procedures such as X-ray and MRI, as well as diagnoses required for surgical reimbursement, were almost perfectly correlated with surgery. These factors were effectively bundled with surgical treatment.

### Health care resource use (HCRU)

We measured HCRU by interactions with the healthcare system, i.e. claims for healthcare encounters in the large commercial data (cumulative count data where appropriate). HCRU was captured longitudinally from the day after the procedure end date until 12, 24, 36, and 48 months after the procedure start date. Encounter types included: outpatient (non-physical therapy) visits, outpatient physical therapy, radiology, surgical procedures, subsequent BMAC or PRP procedures, specialized care for wound healing, emergency or inpatient care, skilled nursing, inpatient rehabilitation, home health services, and opioid medications. Subsequent HCRU encounters were linked to the index procedure using relevant ICD-10 codes. Details of HCRU attribution and episode definitions are in Supplementary Appendix 1.

### Costs

The primary outcome was cost at 12- and 24-months post-procedure. Due to low sample size of patients with extended commercial payer coverage, we assessed cost outcomes out to 36- and 48-months post-procedure as an exploratory aim. We obtained cost estimates for each type of health encounter under two payment models: Medicare and private payer. Medicare costs were estimated using previously published values from external sources (See Supplementary Appendix 1). To estimate private payer costs, we applied three methods. First, Medicare-based costs were multiplied by 2, which is an approximate conversion of Medicare to private payer costs based on an average of multipliers across different cost settings. Second, we used the available private payer cost data and computed average allowed amounts per episode of care (day for outpatient services, and full episode of care for inpatient/emergency). Each HCRU was assigned a cost value, based on this average (See Supplementary Appendix 1 for table of assigned costs). Finally, we summed all eligible private payer costs of those patients who were compared in the matched analysis. This last approach introduced some outlying costs that appeared likely to be data errors. We therefore added a truncated version of this approach by truncating the costs at the 5^th^ and 95^th^ percentile prior to making comparisons.

### Statistical analysis

We first characterized the starting population of eligible surgical and PRP patients with respect to all available covariates, reporting frequency counts and percentages for categorical variables and mean, standard deviation, median, interquartile range, minimum and maximum for continuous variables.

To estimate the probability of receiving orthobiologic treatment, we fit a logistic regression model for all individuals in the comparison set. For each patient, the linear predictor (X*β) was estimated by applying the estimated coefficients from the propensity score model to their individual covariates. We conducted 1:20 greedy matching with respect to the linear predictor, beginning with each patient receiving orthobiologic treatment and seeking 20 matching surgical patients within a caliper of 0.20 times the standard deviation of X*β. Continuous covariates were assessed for non-linearity using restricted cubic splines if any imbalance suggested model mis-specification.

To assess the adequacy of the treatment selection model, we compared baseline characteristics between groups after matching. For all potential confounders, absolute standardized mean differences (ASMDs) were calculated and visualized using standard ASMD plots. ASMDs below 0.10 indicate good balance and below 0.20 moderate balance.

At each timepoint we described HCRU outcomes between treatment comparator groups in the propensity matched population. P-values are included for the comparison of HCRU outcomes, but our purpose was primary descriptive and not hypothesis driven. Finally, we compared aggregate costs at the primary, 12 and 24 month, and exploratory, 36 and 48 month, endpoints between treatment comparator groups in the propensity matched population. The distribution of cost across patients was reported by comparator group and the difference in mean costs computed along with a 95% confidence interval for the difference.

Due to requirements for privacy protection, cell counts less than 11 were suppressed and are reported as <11. To overcome the limitation of cell suppression, we removed the requirement for continuous coverage, at least one outpatient claim, and a relevant orthopedic diagnosis in the 12 months prior to the procedure. This doubled the sample size of PRP patients and allowed for a narrower bound or specific count of some rare events. However, without detailed medical history it was not possible to establish comparability between these patients and a particular surgery. We therefore view this as exploratory.

## RESULTS

After applying eligibility criteria and prior to matching, we identified 262 patients that received PRP injection and 82,152 patients that underwent spine surgery (Supplementary Tables 1-3). After matching, the sample included 133 patients that received PRP matched to 2,560 patients that underwent fusion and 198 patients that received PRP injection matched to 3,960 patients that underwent an LFDF procedure (Tables 1 & 2).

**Table 1.** Post-matching demographic and clinical characteristics – PRP versus fusion.

| Characteristic | PRP<br>N = 128 | Fusion<br>N = 2,560 | ASMD |
| --- | --- | --- | --- |
| Continuous commercial health plan enrollment after index procedure |  |  |  |
| CE for 12 months | 128 (100%) | 2,560 (100%) | 0.000 |
| CE for 24 months | 71 (55%) | 1,737 (68%) | 0.257 |
| CE for 36 months | 41 (32%) | 1,150 (45%) | 0.267 |
| CE for 48 months | 22 (17%) | 724 (28%) | 0.267 |
| Sex |  |  | 0.130 |
| Female | 51 (40%) | 1,185 (46%) |  |
| Male | 77 (60%) | 1,375 (54%) |  |
| Age* |  |  | 0.021 |
| Mean (SD) | 52.1 (11.5) | 52.3 (9.9) |  |
| Median (Q1, Q3) | 55.0 (46.0, 60.0) | 54.0 (46.0, 60.0) |  |
| Geographic region* |  |  | 0.106 |
| Northeast | 19 (15%) | 481 (19%) |  |
| Midwest | 52 (41%) | 1,074 (42%) |  |
| South | 39 (30%) | 703 (27%) |  |
| West | 18 (14%) | 302 (12%) |  |
| Other | <11 | <11 |  |
| Conditions of the lumbar spine compatible with DDD within 12 months prior to the procedure |  |  |  |
| Lumbar disc degeneration or spondylosis* | 102 (80%) | 2,088 (82%) | 0.047 |
| Lumbar spondylolisthesis* | 36 (28%) | 772 (30%) | 0.045 |
| Lumbar stenosis or foraminal narrowing* | 76 (59%) | 1,503 (59%) | 0.014 |
| Lumbar myelopathy* | <11 | 42 (1.6%) | 0.006 |
| Lumbar radiculopathy* | 101 (79%) | 2,117 (83%) | 0.096 |
| Conditions of the lumbar spine not indicative of DDD within 12 months prior to the procedure |  |  |  |
| Herniated disc/intervertebral disc displacement* | 58 (45%) | 1,168 (46%) | 0.006 |
| Other lumbar/lumbosacral disc disorders | <11 | 64 (2.5%) | 0.080 |
| Low back pain | 95 (74%) | 1,928 (75%) | 0.025 |
| Conditions related to the spine (not specific to lumbar) within 12 months prior to the procedure |  |  |  |
| Deforming dorsopathies* | 12 (9.4%) | 206 (8.0%) | 0.047 |
| Kyphosis | <11 | 49 (1.9%) | 0.098 |
| Multi-section conditions compatible with DDD within 12 months prior to the procedure |  |  |  |
| Disc degeneration | 77 (60%) | 1,457 (57%) | 0.066 |
| Disc degeneration - number of regions* |  |  | 0.066 |
| One region | 62 (48%) | 1,170 (46%) |  |
| Two regions | 15 (12%) | 283 (11%) |  |
| Three or more regions | <11 | <11 |  |
| None | 51 (40%) | 1,103 (43%) |  |
| Instability | 35 (27%) | 760 (30%) | 0.052 |
| Instability - number of regions* |  |  | 0.052 |
| One region | 28 (22%) | 597 (23%) |  |
| Two regions | <11 | 144 (5.6%) |  |
| Three or more regions | <11 | 19 (0.7%) |  |
| None | 93 (73%) | 1,800 (70%) |  |
| Spinal stenosis | 71 (55%) | 1,500 (59%) | 0.063 |
| Spinal stenosis - number of regions |  |  | 0.089 |
| One region | 59 (46%) | 1,183 (46%) |  |
| Two regions | 11 (8.6%) | 288 (11%) |  |
| Three or more regions | <11 | 29 (1.1%) |  |
| None | 57 (45%) | 1,060 (41%) |  |
| Spondylosis | 83 (65%) | 1,649 (64%) | 0.009 |
| Spondylosis - number of regions |  |  | 0.024 |
| One region | 55 (43%) | 1,107 (43%) |  |
| Two regions | 25 (20%) | 476 (19%) |  |
| Three or more regions | <11 | 66 (2.6%) |  |
| None | 45 (35%) | 911 (36%) |  |
| Radiculopathy | 97 (76%) | 2,119 (83%) | 0.173 |
| Radiculopathy - number of regions |  |  | 0.174 |
| One region | 60 (47%) | 1,253 (49%) |  |
| Two regions | 27 (21%) | 732 (29%) |  |
| Three or more regions | <11 | 134 (5.2%) |  |
| None | 31 (24%) | 441 (17%) |  |
| Myelopathy | <11 | 159 (6.2%) | 0.147 |
| Myelopathy - number of regions |  |  | 0.147 |
| One region | <11 | 147 (5.7%) |  |
| Two regions | <11 | 11 (0.4%) |  |
| Three or more regions | <11 | <11 |  |
| None | 124 (97%) | 2,401 (94%) |  |
| Musculoskeletal conditions (not exclusive to the spine) within 12 months prior to the procedure |  |  |  |
| Poor bone quality | <11 | 60 (2.3%) | 0.056 |
| Osteoarthritis in other parts of the body | 34 (27%) | 495 (19%) | 0.172 |
| General comorbidity diagnoses within 12 months prior to the procedure |  |  |  |
| Obesity* | 22 (17%) | 501 (20%) | 0.062 |
| Diabetes* | 14 (11%) | 279 (11%) | 0.001 |
| Depression/Anxiety/Bipolar* | 34 (27%) | 634 (25%) | 0.041 |
| Medications within 12 months prior to the procedure |  |  |  |
| Opioid or tramadol medication* | 46 (36%) | 820 (32%) | 0.083 |
| NSAID medication* | 38 (30%) | 690 (27%) | 0.061 |
| Neuropathic pain medication* | 26 (20%) | 549 (21%) | 0.028 |
| Intervention history in the 12 months prior to the procedure |  |  |  |
| Diagnostic or therapeutic injection (CPT 62322, 62323) * | 14 (11%) | 243 (9.5%) | 0.048 |
| Injection or anesthetic or steroid (CPT 64483, 64484) * | 26 (20%) | 471 (18%) | 0.048 |
| Oral corticosteroids | 38 (30%) | 816 (32%) | 0.047 |
| Physical therapy | 64 (50%) | 1,231 (48%) | 0.038 |
| Greater than 6 PT visits | 33 (26%) | 630 (25%) | 0.027 |
| Number of PT visits* |  |  | 0.051 |
| Mean (SD) | 5.4 (9.3) | 4.9 (9.2) |  |
| Median (Q1, Q3) | 0.5 (0.0, 7.0) | 0.0 (0.0, 6.0) |  |
| Health seeking behavior in the 12 months prior to the procedure |  |  |  |
| Number of outpatient visits |  |  | 0.000 |
| Mean (SD) | 23.1 (16.9) | 23.1 (15.7) |  |
| Median (Q1, Q3) | 19.0 (12.0, 30.0) | 19.0 (12.0, 29.0) |  |
| ER visit* | 22 (17%) | 378 (15%) | 0.066 |
| Number of ER visits |  |  | 0.004 |
| Mean (SD) | 0.2 (0.6) | 0.2 (0.7) |  |
| Median (Q1, Q3) | 0.0 (0.0, 0.0) | 0.0 (0.0, 0.0) |  |
| Chiropractic visit* | 20 (16%) | 543 (21%) | 0.144 |
| Number of chiropractic visit |  |  | 0.018 |
| Mean (SD) | 1.9 (6.2) | 2.0 (6.2) |  |
| Median (Q1, Q3) | 0.0 (0.0, 0.0) | 0.0 (0.0, 0.0) |  |
| Spine x-ray | 72 (56%) | 1,929 (75%) | N/A |
| Spine MRI | 99 (77%) | 2,231 (87%) | N/A |
| Spine x-ray or spine MRI | 108 (84%) | 2,407 (94%) | N/A |
| Other severity proxies in the 12 months prior to the procedure |  |  |  |
| Spinal stenosis outpatient visit | 71 (55%) | 1,483 (58%) | 0.050 |
| Number of spinal stenosis outpatient visits* |  |  | 0.003 |
| Mean (SD) | 1.3 (2.3) | 1.3 (2.0) |  |
| Median (Q1, Q3) | 1.0 (0.0, 1.0) | 1.0 (0.0, 2.0) |  |
| Disc degeneration outpatient visit | 77 (60%) | 1,451 (57%) | 0.071 |
| Number of disc degeneration outpatient visits |  |  | 0.054 |
| Mean (SD) | 1.9 (4.5) | 1.7 (3.2) |  |
| Median (Q1, Q3) | 1.0 (0.0, 2.0) | 1.0 (0.0, 2.0) |  |
| Instability outpatient visit | 35 (27%) | 757 (30%) | 0.049 |
| Number of instability outpatient visits* |  |  | 0.040 |
| Mean (SD) | 0.7 (2.4) | 0.8 (1.8) |  |
| Median (Q1, Q3) | 0.0 (0.0, 1.0) | 0.0 (0.0, 1.0) |  |
| Osteoarthritis outpatient visit | 34 (27%) | 486 (19%) | 0.181 |
| Number of osteoarthritis outpatient visits |  |  | 0.097 |
| Mean (SD) | 0.8 (2.2) | 0.6 (2.4) |  |
| Median (Q1, Q3) | 0.0 (0.0, 1.0) | 0.0 (0.0, 0.0) |  |
| Less invasive intervention indicator (CPT 62322, 62323, 64483, 64484, GT 6 PT visits) * | 60 (47%) | 1,069 (42%) | 0.103 |

\*Indicates variables used in propensity score matching
CE, continuous enrollment; DDD, degenerative disc disease; SD, standard deviation; CPT, current procedural terminology; MRI, magnetic resonance imaging; PT, physical therapy; N/A where ASMDs are not computed for procedures that are likely part of pre-op for surgery

**Table 2.** Post-matching demographic and clinical characteristics – PRP versus LFDF.

| Characteristic | PRP<br>N = 198 | LFDF<br>N = 3,960 | ASMD |
| --- | --- | --- | --- |
| Continuous commercial health plan enrollment after index procedure |  |  |  |
| CE for 12 months | 198 (100%) | 3,960 (100%) | 0.000 |
| CE for 24 months | 116 (59%) | 2,809 (71%) | 0.261 |
| CE for 36 months | 77 (39%) | 1,953 (49%) | 0.211 |
| CE for 48 months | 50 (25%) | 1,271 (32%) | 0.152 |
| Sex |  |  | 0.048 |
| Female | 79 (40%) | 1,488 (38%) |  |
| Male | 119 (60%) | 2,472 (62%) |  |
| Age* |  |  | 0.039 |
| Mean (SD) | 50.4 (12.1) | 50.0 (11.7) |  |
| Median (Q1, Q3) | 53.0 (43.0, 60.0) | 52.0 (41.0, 59.0) |  |
| Geographic region* |  |  | 0.083 |
| Northeast | 39 (20%) | 815 (21%) |  |
| Midwest | 75 (38%) | 1,609 (41%) |  |
| South | 49 (25%) | 956 (24%) |  |
| West | 35 (18%) | 579 (15%) |  |
| Other | <11 | <11 |  |
| Conditions of the lumbar spine compatible with DDD within 12 months prior to the procedure |  |  |  |
| Lumbar disc degeneration or spondylosis* | 147 (74%) | 3,113 (79%) | 0.103 |
| Lumbar spondylolisthesis* | 36 (18%) | 604 (15%) | 0.079 |
| Lumbar stenosis or foraminal narrowing* | 92 (46%) | 1,805 (46%) | 0.018 |
| Lumbar myelopathy* | <11 | 43 (1.1%) | 0.007 |
| Lumbar radiculopathy* | 168 (85%) | 3,257 (82%) | 0.070 |
| Conditions of the lumbar spine not indicative of DDD within 12 months prior to the procedure |  |  |  |
| Herniated disc/intervertebral disc displacement* | 92 (46%) | 1,869 (47%) | 0.015 |
| Other lumbar/lumbosacral disc disorders | <11 | 110 (2.8%) | 0.015 |
| Low back pain | 139 (70%) | 2,763 (70%) | 0.009 |
| Conditions related to the spine (not specific to lumbar) within 12 months prior to the procedure |  |  |  |
| Deforming dorsopathies* | <11 | 180 (4.5%) | 0.024 |
| Kyphosis | <11 | 34 (0.9%) | 0.043 |
| Multi-section conditions compatible with DDD within 12 months prior to the procedure |  |  |  |
| Disc degeneration | 101 (51%) | 2,082 (53%) | 0.031 |
| Disc degeneration - number of regions* |  |  | 0.055 |
| One region | 85 (43%) | 1,808 (46%) |  |
| Two regions | 16 (8.1%) | 268 (6.8%) |  |
| Three or more regions | <11 | <11 |  |
| None | 97 (49%) | 1,878 (47%) |  |
| Instability | 32 (16%) | 553 (14%) | 0.061 |
| Instability - number of regions* |  |  | 0.061 |
| One region | 27 (14%) | 466 (12%) |  |
| Two regions | <11 | 79 (2.0%) |  |
| Three or more regions | <11 | <11 |  |
| None | 166 (84%) | 3,407 (86%) |  |
| Spinal stenosis | 75 (38%) | 1,629 (41%) | 0.067 |
| Spinal stenosis - number of regions |  |  | 0.067 |
| One region | 62 (31%) | 1,355 (34%) |  |
| Two regions | 12 (6.1%) | 247 (6.2%) |  |
| Three or more regions | <11 | 27 (0.7%) |  |
| None | 123 (62%) | 2,331 (59%) |  |
| Spondylosis | 113 (57%) | 2,073 (52%) | 0.095 |
| Spondylosis - number of regions |  |  | 0.187 |
| One region | 77 (39%) | 1,594 (40%) |  |
| Two regions | 33 (17%) | 408 (10%) |  |
| Three or more regions | <11 | 71 (1.8%) |  |
| None | 85 (43%) | 1,887 (48%) |  |
| Radiculopathy | 163 (82%) | 3,167 (80%) | 0.060 |
| Radiculopathy - number of regions |  |  | 0.148 |
| One region | 105 (53%) | 1,807 (46%) |  |
| Two regions | 47 (24%) | 1,167 (29%) |  |
| Three or more regions | 11 (5.6%) | 193 (4.9%) |  |
| None | 35 (18%) | 793 (20%) |  |
| Myelopathy | <11 | 86 (2.2%) | 0.011 |
| Myelopathy - number of regions |  |  | 0.039 |
| One region | <11 | 83 (2.1%) |  |
| Two regions | <11 | <11 |  |
| Three or more regions | <11 | <11 |  |
| None | 194 (98%) | 3,874 (98%) |  |
| Musculoskeletal conditions (not exclusive to the spine) within 12 months prior to the procedure |  |  |  |
| Poor bone quality | <11 | 51 (1.3%) | 0.026 |
| Osteoarthritis in other parts of the body | 45 (23%) | 541 (14%) | 0.237 |
| General comorbidity diagnoses within 12 months prior to the procedure |  |  |  |
| Obesity* | 26 (13%) | 537 (14%) | 0.013 |
| Diabetes* | 15 (7.6%) | 297 (7.5%) | 0.003 |
| Depression/Anxiety/Bipolar* | 39 (20%) | 823 (21%) | 0.027 |
| Medications within 12 months prior to the procedure |  |  |  |
| Opioid or tramadol medication* | 67 (34%) | 1,197 (30%) | 0.077 |
| NSAID medication* | 50 (25%) | 977 (25%) | 0.013 |
| Neuropathic pain medication* | 31 (16%) | 661 (17%) | 0.028 |
| Intervention history in the 12 months prior to the procedure |  |  |  |
| Diagnostic or therapeutic injection (CPT 62322, 62323) * | 13 (6.6%) | 279 (7.0%) | 0.019 |
| Injection or anesthetic or steroid (CPT 64483, 64484) * | 32 (16%) | 698 (18%) | 0.039 |
| Oral corticosteroids | 50 (25%) | 1,070 (27%) | 0.040 |
| Physical therapy | 108 (55%) | 2,085 (53%) | 0.038 |
| Greater than 6 PT visits | 57 (29%) | 1,165 (29%) | 0.014 |
| Number of PT visits* |  |  | 0.004 |
| Mean (SD) | 5.7 (9.0) | 5.8 (9.7) |  |
| Median (Q1, Q3) | 1.0 (0.0, 9.0) | 1.0 (0.0, 8.0) |  |
| Health seeking behavior in the 12 months prior to the procedure |  |  |  |
| Number of outpatient visits |  |  | 0.088 |
| Mean (SD) | 22.0 (14.0) | 20.7 (15.2) |  |
| Median (Q1, Q3) | 19.0 (12.0, 30.0) | 17.0 (10.0, 27.0) |  |
| ER visit* | 33 (17%) | 604 (15%) | 0.039 |
| Number of ER visits |  |  | 0.018 |
| Mean (SD) | 0.2 (0.6) | 0.2 (0.6) |  |
| Median (Q1, Q3) | 0.0 (0.0, 0.0) | 0.0 (0.0, 0.0) |  |
| Chiropractic visit* | 57 (29%) | 1,094 (28%) | 0.026 |
| Number of chiropractic visit |  |  | 0.090 |
| Mean (SD) | 3.4 (7.9) | 2.8 (6.9) |  |
| Median (Q1, Q3) | 0.0 (0.0, 2.0) | 0.0 (0.0, 1.0) |  |
| Spine x-ray | 97 (49%) | 2,737 (69%) | N/A |
| Spine MRI | 146 (74%) | 3,467 (88%) | N/A |
| Spine x-ray or spine MRI | 160 (81%) | 3,671 (93%) | N/A |
| Other severity proxies in the 12 months prior to the procedure |  |  |  |
| Spinal stenosis outpatient visit | 75 (38%) | 1,625 (41%) | 0.065 |
| Number of spinal stenosis outpatient visits* |  |  | 0.032 |
| Mean (SD) | 0.9 (1.9) | 0.9 (2.4) |  |
| Median (Q1, Q3) | 0.0 (0.0, 1.0) | 0.0 (0.0, 1.0) |  |
| Disc degeneration outpatient visit | 101 (51%) | 2,078 (52%) | 0.029 |
| Number of disc degeneration outpatient visits |  |  | 0.077 |
| Mean (SD) | 1.6 (3.9) | 1.3 (3.2) |  |
| Median (Q1, Q3) | 1.0 (0.0, 2.0) | 1.0 (0.0, 1.0) |  |
| Instability outpatient visit | 32 (16%) | 553 (14%) | 0.061 |
| Number of instability outpatient visits* |  |  | 0.018 |
| Mean (SD) | 0.3 (0.9) | 0.3 (1.4) |  |
| Median (Q1, Q3) | 0.0 (0.0, 0.0) | 0.0 (0.0, 0.0) |  |
| Osteoarthritis outpatient visit | 45 (23%) | 538 (14%) | 0.239 |
| Number of osteoarthritis outpatient visits |  |  | 0.210 |
| Mean (SD) | 0.9 (3.2) | 0.3 (1.6) |  |
| Median (Q1, Q3) | 0.0 (0.0, 0.0) | 0.0 (0.0, 0.0) |  |
| Less invasive intervention indicator (CPT 62322, 62323, 64483, 64484, GT 6 PT visits) * | 87 (44%) | 1,736 (44%) | 0.002 |

\*Indicates variables used in propensity score matching
CE, continuous enrollment; DDD, degenerative disc disease; SD, standard deviation; CPT, current procedural terminology; MRI, magnetic resonance imaging; PT, physical therapy; N/A where ASMDs are not computed for procedures that are likely part of pre-op for surgery

In both comparison cohorts, all variables that were included in propensity score matching were well balanced (ASMDs <0.10) as were nearly all measured variables (Tables 1 & 2), except for those that were expected to be bundled with surgery (pre-op X-ray, CT, MRI).

### Health care use

#### PRP versus fusion

Rates of post-procedure fusion and LFDF following PRP were below the cell suppression limit of <11 events at 12 and 24 months among those continuously enrolled during those follow-up timeframes (Table 3 & 4). Rates of re-operation (fusion) and LFDF among those initially receiving fusion were 7.4% and 3.3%, respectively, at 12 months and 9.3% and 4.8%, respectively, at 24 months. Rates of post-procedure PRP injection, i.e., booster, were 14.1% at 12 months and below the cell suppression limit at 24 months.

**Table 3.** Healthcare Resource Utilization 12 months post procedure – PRP versus fusion.

| **Characteristic** | **PRP**<br>**N = 128** | **Fusion**<br>**N = 2,560** | **p-value** |
| --- | --- | --- | --- |
| Number of outpatient visits |  |  | < 0.001 |
| Mean (SD) | 4.6 (5.3) | 5.9 (5.4) |  |
| Median (Q1, Q3) | 3.0 (1.0, 6.0) | 4.0 (2.0, 8.0) |  |
| Number of outpatient physical therapy visits |  |  | 0.25 |
| Mean (SD) | 5.9 (13.7) | 5.8 (10.4) |  |
| Median (Q1, Q3) | 0.0 (0.0, 5.5) | 0.0 (0.0, 8.0) |  |
| Spine x-ray | 29 (22.7%) | 2,385 (93.2%) | < 0.001 |
| MRI of spine | 29 (22.7%) | 561 (21.9%) | 0.93 |
| CT of spine | <11 | 440 (17.2%) | < 0.001 |
| Subsequent fusion | <11 | 190 (7.4%) | 0.02 |
| Subsequent LFDF | <11 | 84 (3.3%) | -- |
| Subsequent BMAC/PRP | 18 (14.1%) | <11 | -- |
| Specialized care for wound healing (inpatient or outpatient) | <11 | 48 (1.9%) | -- |
| Post-procedural complications inpatient care | <11 | 41 (1.6%) | -- |
| Cerebrospinal fluid leak inpatient care | <11 | <11 | -- |
| Mechanical device complications inpatient care | <11 | 11 (0.4%) | -- |
| Hematoma inpatient care | <11 | 12 (0.5%) | -- |
| Wound infection inpatient care | <11 | 35 (1.4%) | -- |
| Sepsis inpatient care | <11 | 17 (0.7%) | -- |
| Deep vein thrombosis inpatient care | <11 | 11 (0.4%) | -- |
| Pulmonary embolism inpatient care | <11 | 16 (0.6%) | -- |
| Other post-surgical embolism inpatient care | <11 | <11 | -- |
| DVT, PT, or OE inpatient care | <11 | 21 (0.8%) | -- |
| SNF services | <11 | <11 | -- |
| IRF services | <11 | <11 | -- |
| Home health services | 18 (14.1%) | 1,029 (40.2%) | < 0.001 |
| Opioid medication | 43 (33.6%) | 1,043 (40.7%) | 0.13 |

SD, standard deviation; MRI, magnetic resonance imaging; CT, computed tomography; LFDF, laminectomy, foraminotomy, discectomy, and facetectomy; BMAC, bone marrow aspirate concentrate; PRP, platelet rich plasma; CSF, cerebrospinal fluid; VDT, deep vein thrombosis; PE, pulmonary embolism; OE, obstructive event; SNF, skilled nursing facility; IRF, inpatient rehabilitation facility; Rx, prescription.

**Table 4.** Healthcare Resource Utilization 24 months post procedure – PRP versus fusion.

| Characteristic | PRP<br>N = 71 | Fusion<br>N = 1,737 | p-value |
| --- | --- | --- | --- |
| Number of outpatient visits |  |  | 0.03 |
| Mean (SD) | 7.8 (9.0) | 9.1 (9.1) |  |
| Median (Q1, Q3) | 5.0 (1.0, 12.0) | 6.0 (3.0, 12.0) |  |
| Number of outpatient physical therapy visits |  |  | 0.69 |
| Mean (SD) | 11.2 (24.5) | 7.3 (13.6) |  |
| Median (Q1, Q3) | 0.0 (0.0, 7.0) | 1.0 (0.0, 10.0) |  |
| Spine x-ray | 21 (29.6%) | 1,641 (94.5%) | < 0.001 |
| MRI of spine | 22 (31.0%) | 604 (34.8%) | 0.6 |
| CT of spine | <11 | 415 (23.9%) | 0.004 |
| Subsequent fusion | <11 | 162 (9.3%) | 0.04 |
| Subsequent LFDF | <11 | 84 (4.8%) | -- |
| Subsequent BMAC/PRP | <11 | <11 | -- |
| Specialized care for wound healing (inpatient or outpatient) | <11 | 39 (2.2%) | -- |
| Post-procedural complications inpatient care | <11 | 43 (2.5%) | -- |
| Cerebrospinal fluid leak inpatient care | <11 | <11 | -- |
| Mechanical device complications inpatient care | <11 | 11 (0.6%) | -- |
| Hematoma inpatient care | <11 | <11 | -- |
| Wound infection inpatient care | <11 | 27 (1.6%) | -- |
| Sepsis inpatient care | <11 | 20 (1.2%) | -- |
| Deep vein thrombosis inpatient care | <11 | <11 | -- |
| Pulmonary embolism inpatient care | <11 | 16 (0.9%) | -- |
| Other post-surgical embolism inpatient care | <11 | <11 | -- |
| DVT, PT, or OE inpatient care | <11 | 19 (1.1%) | -- |
| SNF services | <11 | <11 | -- |
| IRF services | <11 | <11 | -- |
| Home health services | 15 (21.1%) | 795 (45.8%) | < 0.001 |
| Opioid medication | 28 (39.4%) | 778 (44.8%) | 0.44 |

SD, standard deviation; MRI, magnetic resonance imaging; CT, computed tomography; LFDF, laminectomy, foraminotomy, discectomy, and facetectomy; BMAC, bone marrow aspirate concentrate; PRP, platelet rich plasma; CSF, cerebrospinal fluid; VDT, deep vein thrombosis; PE, pulmonary embolism; OE, obstructive event; SNF, skilled nursing facility; IRF, inpatient rehabilitation facility; Rx, prescription.

At 12 months, we observed significantly higher rates of post-procedure radiographs (93.2% vs 22.7%), home health service use (40.2% vs 14.1%), and outpatient visits (5.9 vs 4.6) among those undergoing fusion. Rates of opioid use, MRI, and physical therapy visits were not significantly different. At 24 months, we observed higher rates of spine radiographs (94.5% vs 29.6%), home health services (45.8% vs 21.1%), and outpatient visits (9.1 vs 7.8) for the those that underwent fusion, with no group differences in opioid use, MRI, and physical therapy visits.

#### PRP versus LFDF

Rates of LFDF and fusion following PRP were below the cell suppression limit of <11 events at 12 and 24 months among those continuously enrolled during those follow-up timeframes (Tables 5 & 6). Rates of re-operation (LFDF) and fusion among those initially receiving LFDF were 21.8% and 3.1%, respectively, at 12 months and 23.7% and 5.3%, respectively, at 24 months. Rates of post-procedure PRP injection, i.e., booster, were 11.6% at 12 months and 12.1% at 24 months.

**Table 5:** Healthcare Resource Utilization 12 months post procedure – PRP versus LFDF.

| Characteristic | PRP<br>N = 198 | LFDF<br>N = 3,960 | p-value |
| --- | --- | --- | --- |
| Number of outpatient visits |  |  | < 0.001 |
| Mean (SD) | 4.5 (5.6) | 3.2 (4.3) |  |
| Median (Q1, Q3) | 3.0 (1.0, 6.0) | 2.0 (0.0, 4.0) |  |
| Number of outpatient physical therapy visits |  |  | 0.5 |
| Mean (SD) | 5.1 (12.3) | 4.7 (9.1) |  |
| Median (Q1, Q3) | 0.0 (0.0, 5.0) | 0.0 (0.0, 6.0) |  |
| Spine x-ray | 39 (19.7%) | 1,344 (33.9%) | < 0.001 |
| MRI of spine | 36 (18.2%) | 949 (24.0%) | 0.07 |
| CT of spine | <11 | 151 (3.8%) | 0.71 |
| Subsequent fusion | <11 | 124 (3.1%) | 0.28 |
| Subsequent LFDF | <11 | 862 (21.8%) | < 0.001 |
| Subsequent BMAC/PRP | 23 (11.6%) | <11 | -- |
| Specialized care for wound healing (inpatient or outpatient) | <11 | 48 (1.2%) | -- |
| Post-procedural complications inpatient care | <11 | 44 (1.1%) | -- |
| Cerebrospinal fluid leak inpatient care | <11 | 21 (0.5%) | -- |
| Mechanical device complications inpatient care | <11 | <11 | -- |
| Hematoma inpatient care | <11 | 12 (0.3%) | -- |
| Wound infection inpatient care | <11 | 35 (0.9%) | -- |
| Sepsis inpatient care | <11 | 26 (0.7%) | -- |
| Deep vein thrombosis inpatient care | <11 | 13 (0.3%) | -- |
| Pulmonary embolism inpatient care | <11 | 13 (0.3%) | -- |
| Other post-surgical embolism inpatient care | <11 | <11 | -- |
| DVT, PT, or OE inpatient care | <11 | 20 (0.5%) | -- |
| SNF services | <11 | <11 | -- |
| IRF services | <11 | <11 | -- |
| Home health services | 23 (11.6%) | 724 (18.3%) | 0.02 |
| Opioid medication | 69 (34.8%) | 1,158 (29.2%) | 0.11 |

SD, standard deviation; MRI, magnetic resonance imaging; CT, computed tomography; LFDF, laminectomy, foraminotomy, discectomy, and facetectomy; BMAC, bone marrow aspirate concentrate; PRP, platelet rich plasma; IP, inpatient; CSF, cerebrospinal fluid; VDT, deep vein thrombosis; PE, pulmonary embolism; OE, obstructive event; SNF, skilled nursing facility; IRF, inpatient rehabilitation facility; Rx, prescription.

**Table 6.** Healthcare Resource Utilization 24 months post procedure – PRP versus LFDF.

| Characteristic | PRP<br>N = 116 | LFDF<br>N = 2,809 | p-value |
| --- | --- | --- | --- |
| Number of outpatient visits |  |  | < 0.001 |
| Mean (SD) | 7.9 (9.3) | 5.3 (7.1) |  |
| Median (Q1, Q3) | 5.0 (2.0, 11.0) | 3.0 (1.0, 7.0) |  |
| Number of outpatient physical therapy visits |  |  | 0.8 |
| Mean (SD) | 9.4 (20.6) | 6.2 (12.4) |  |
| Median (Q1, Q3) | 0.0 (0.0, 7.5) | 0.0 (0.0, 7.0) |  |
| Spine x-ray | 27 (23.3%) | 1,157 (41.2%) | < 0.001 |
| MRI of spine | 32 (27.6%) | 924 (32.9%) | 0.27 |
| CT of spine | <11 | 183 (6.5%) | 0.99 |
| Subsequent fusion | <11 | 149 (5.3%) | 0.14 |
| Subsequent LFDF | <11 | 667 (23.7%) | < 0.001 |
| Subsequent Regenxx SCP | 14 (12.1%) | <11 | -- |
| Specialized care for wound healing (inpatient or outpatient) | <11 | 46 (1.6%) | -- |
| Post-procedural complications inpatient care | <11 | 42 (1.5%) | -- |
| Cerebrospinal fluid leak inpatient care | <11 | 17 (0.6%) | -- |
| Mechanical device complications inpatient care | <11 | <11 | -- |
| Hematoma inpatient care | <11 | <11 | -- |
| Wound infection inpatient care | <11 | 33 (1.2%) | -- |
| Sepsis inpatient care | <11 | 24 (0.9%) | -- |
| Deep vein thrombosis inpatient care | <11 | 15 (0.5%) | -- |
| Pulmonary embolism inpatient care | <11 | 12 (0.4%) | -- |
| Other post-surgical embolism inpatient care | <11 | <11 | -- |
| DVT, PT, or OE inpatient care | <11 | 21 (0.7%) | -- |
| SNF services | <11 | <11 | -- |
| IRF services | <11 | <11 | -- |
| Home health services | 22 (19.0%) | 697 (24.8%) | 0.19 |
| Opioid medication | 48 (41.4%) | 960 (34.2%) | 0.13 |

SD, standard deviation; MRI, magnetic resonance imaging; CT, computed tomography; LFDF, laminectomy, foraminotomy, discectomy, and facetectomy; BMAC, bone marrow aspirate concentrate; PRP, platelet rich plasma; IP, inpatient; CSF, cerebrospinal fluid; VDT, deep vein thrombosis; PE, pulmonary embolism; OE, obstructive event; SNF, skilled nursing facility; IRF, inpatient rehabilitation facility; Rx, prescription.

At 12 months, we observed higher rates of post-procedure radiographs (33.9% vs 19.7%) and home health service use (18.3% vs 11.6%), but lower rates of outpatient visits (3.2 vs 4.5) among those that underwent LFDF. Rates of opioid use, MRI, and physical therapy visits were not significantly different. At 24 months, we observed lower rates of outpatient visits (5.3 vs 7.9), but higher rates of post-procedure radiographs (41.2% vs 23.3%) among those that underwent LFDF. Rates of all other types of health care use were not different between groups.

### Costs

#### PRP versus fusion

At 12 and 24 months, estimated mean costs were significantly higher among those undergoing fusion compared to fusion, with mean cost difference point estimates ranging from $28,454 to $66,756 (Table 7).

**Table 7.**
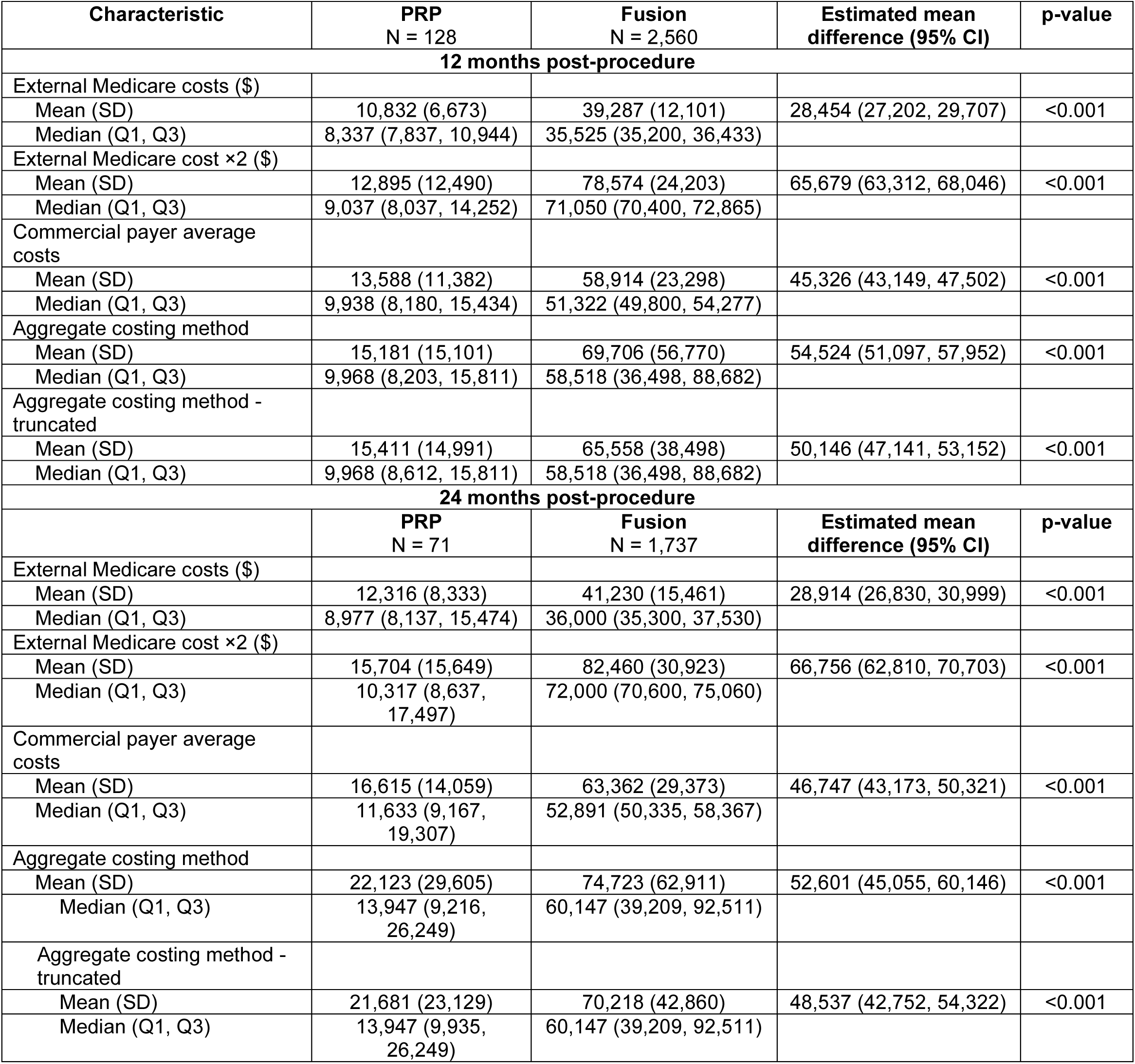
Costs – PRP versus fusion at 12- and 24-months post procedure.

| Characteristic | PRP<br>N = 128 | Fusion<br>N = 2,560 | Estimated mean<br>difference (95% CI) | p-value |
| --- | --- | --- | --- | --- |
| **12 months post-procedure** |  |  |  |  |
| External Medicare costs (\$) | | | | |
| Mean (SD) | 10,832 (6,673) | 39,287 (12,101) | 28,454 (27,202, 29,707) | <0.001 |
| Median (Q1, Q3) | 8,337 (7,837, 10,944) | 35,525 (35,200, 36,433) |  |  |
| External Medicare cost ×2 (\$) | | | | |
| Mean (SD) | 12,895 (12,490) | 78,574 (24,203) | 65,679 (63,312, 68,046) | <0.001 |
| Median (Q1, Q3) | 9,037 (8,037, 14,252) | 71,050 (70,400, 72,865) |  |  |
| Commercial payer average costs |  |  |  |  |
| Mean (SD) | 13,588 (11,382) | 58,914 (23,298) | 45,326 (43,149, 47,502) | <0.001 |
| Median (Q1, Q3) | 9,938 (8,180, 15,434) | 51,322 (49,800, 54,277) |  |  |
| Aggregate costing method |  |  |  |  |
| Mean (SD) | 15,181 (15,101) | 69,706 (56,770) | 54,524 (51,097, 57,952) | <0.001 |
| Median (Q1, Q3) | 9,968 (8,203, 15,811) | 58,518 (36,498, 88,682) |  |  |
| Aggregate costing method - truncated |  |  |  |  |
| Mean (SD) | 15,411 (14,991) | 65,558 (38,498) | 50,146 (47,141, 53,152) | <0.001 |
| Median (Q1, Q3) | 9,968 (8,612, 15,811) | 58,518 (36,498, 88,682) |  |  |
| **24 months post-procedure** |  |  |  |  |
|  | PRP<br>N = 71 | Fusion<br>N = 1,737 | Estimated mean<br>difference (95% CI) | p-value |
| External Medicare costs (\$) | | | | |
| Mean (SD) | 12,316 (8,333) | 41,230 (15,461) | 28,914 (26,830, 30,999) | <0.001 |
| Median (Q1, Q3) | 8,977 (8,137, 15,474) | 36,000 (35,300, 37,530) |  |  |
| External Medicare cost ×2 (\$) | | | | |
| Mean (SD) | 15,704 (15,649) | 82,460 (30,923) | 66,756 (62,810, 70,703) | <0.001 |
| Median (Q1, Q3) | 10,317 (8,637, 17,497) | 72,000 (70,600, 75,060) |  |  |
| Commercial payer average costs |  |  |  |  |
| Mean (SD) | 16,615 (14,059) | 63,362 (29,373) | 46,747 (43,173, 50,321) | <0.001 |
| Median (Q1, Q3) | 11,633 (9,167, 19,307) | 52,891 (50,335, 58,367) |  |  |
| Aggregate costing method |  |  |  |  |
| Mean (SD) | 22,123 (29,605) | 74,723 (62,911) | 52,601 (45,055, 60,146) | <0.001 |
| Median (Q1, Q3) | 13,947 (9,216, 26,249) | 60,147 (39,209, 92,511) |  |  |
| Aggregate costing method - truncated |  |  |  |  |
| Mean (SD) | 21,681 (23,129) | 70,218 (42,860) | 48,537 (42,752, 54,322) | <0.001 |
| Median (Q1, Q3) | 13,947 (9,935, 26,249) | 60,147 (39,209, 92,511) |  |  |

#### PRP versus LFDF

At 12 and 24 months, estimated mean costs were significantly higher among those undergoing LFDF, except when considering the aggregate costing method at 24 months (mean difference = 4,447 (−687, 9,582). Mean cost difference point estimates ranged from $2,579 to $17,944 at 12 months and from $2,488 to $17,931 at 24 months. (Table 8).

**Table 8.** Costs – PRP versus LFDF at 12- and 24-months post procedure.

| Characteristic | PRP<br>N = 198 | LFDF<br>N = 3,960 | Estimated mean<br>difference (95% CI) | p-value |
| --- | --- | --- | --- | --- |
| **12 months post-procedure** |  |  |  |  |
| External Medicare costs (\$) | | | | |
| Mean (SD) | 10,492 (6,310) | 13,071 (9,528) | 2,579 (1,649, 3,509) | <0.001 |
| Median (Q1, Q3) | 8,237 (7,737, 9,877) | 9,300 (9,000, 12,253) |  |  |
| External Medicare cost ×2 (\$) | | | | |
| Mean (SD) | 12,383 (11,785) | 26,142 (19,056) | 13,758 (12,008, 15,508) | <0.001 |
| Median (Q1, Q3) | 8,837 (7,837, 12,117) | 18,600 (18,000, 24,505) |  |  |
| Commercial payer average costs |  |  |  |  |
| Mean (SD) | 12,998 (10,735) | 30,942 (20,072) | 17,944 (16,319, 19,569) | <0.001 |
| Median (Q1, Q3) | 9,625 (8,081, 13,343) | 22,459 (21,024, 37,495) |  |  |
| Aggregate costing method |  |  |  |  |
| Mean (SD) | 14,140 (13,866) | 20,697 (32,316) | 6,558 (4,375, 8,740) | <0.001 |
| Median (Q1, Q3) | 9,334 (7,937, 15,274) | 14,440 (9,786, 22,096) |  |  |
| Aggregate costing method - truncated |  |  |  |  |
| Mean (SD) | 13,435 (9,998) | 18,316 (12,740) | 4,881 (3,429, 6,333) | <0.001 |
| Median (Q1, Q3) | 9,334 (7,937, 15,274) | 14,440 (9,786, 22,096) |  |  |
| **24 months post-procedure** |  |  |  |  |
|  | PRP<br>N = 116 | LFDF<br>N = 2,809 | Estimated mean<br>difference (95% CI) | p-value |
| External Medicare costs (\$) | | | | |
| Mean (SD) | 12,291 (9,710) | 14,779 (12,511) | 2,488 (653, 4,323) | 0.008 |
| Median (Q1, Q3) | 8,937 (8,037, 12,622) | 9,700 (9,100, 18,000) |  |  |
| External Medicare cost ×2 (\$) | | | | |
| Mean (SD) | 15,891 (18,786) | 29,558 (25,022) | 13,667 (10,109, 17,224) | <0.001 |
| Median (Q1, Q3) | 10,237 (8,437, 16,142) | 19,400 (18,200, 36,000) |  |  |
| Commercial payer average costs |  |  |  |  |
| Mean (SD) | 16,737 (16,113) | 34,667 (24,998) | 17,931 (14,843, 21,019) | <0.001 |
| Median (Q1, Q3) | 11,662 (8,969, 16,913) | 23,729 (21,341, 42,048) |  |  |
| Aggregate costing method |  |  |  |  |
| Mean (SD) | 19,905 (27,168) | 24,353 (35,072) | 4,447 (-687, 9,582) | 0.09 |
| Median (Q1, Q3) | 11,077 (8,732, 17,921) | 15,578 (10,266, 24,917) |  |  |
| Aggregate costing method - truncated |  |  |  |  |
| Mean (SD) | 17,783 (15,525) | 21,491 (17,551) | 3,709 (797, 6,621) | 0.013 |
| Median (Q1, Q3) | 11,077 (8,732, 17,921) | 15,578 (10,266, 24,917) |  |  |

### Exploratory analyses at 36 and 48 months

In the fusion comparator analysis, rates of post-procedure fusion and LFDF for those receiving PRP were below the cell suppression limit of <11 events at 36 and 48 months among those continuously enrolled during those follow-up timeframes.

Rates of spine radiographs and home health services continued to be higher in the surgical cohort at 36 and 48 months (Supplementary Tables 4 & 5). At 36 and 48 months, costs were higher among those that underwent fusion using all costing methods (Supplementary Table 6).

In the LFDF comparator analysis, rates of post-procedure fusion and LFDF for those receiving PRP were below the cell suppression limit of <11 events at 36 and 48 months among those continuously enrolled during those follow-up timeframes.

At 36 and 48 months, rates of physical therapy use were higher in those who received PRP, while rates of spine radiographs were higher in those who underwent surgery (Supplementary Tables 7 & 8). At 36 and 48 months, costs were consistently higher in the LFDF group but only when using external costs, external inflated costs, and commercial payer costing methods. Costs were generally not different between groups when using the aggregate costing methods (Supplementary Table 9).

### Expanded Analysis

The analysis with reduced exclusion criteria to overcome cell suppression included n=540 undergoing PRP, n=41,927 undergoing fusion, and n=86,163 undergoing LFDF (Supplementary Tables 10 & 11). Among those who received PRP, subsequent fusion and LFDF remained below the cell suppression limit through 48 months, except for a rate of 3.8% (n=13) receiving LFDF at 24 months (Supplementary Tables 12-19). Rates of PRP re-injections ranged from 9.7% to 12.4% across all follow-ups.

## DISCUSSION

This analysis suggests PRP may offer a cost-efficient alternative to spine surgery for select patients with degenerative spinal conditions. Subsequent health care costs were substantially lower among those receiving PRP compared to surgery, primarily reflecting the large cost differential between index procedures, and higher rates of reoperation and imaging in the surgical cohort. Importantly, these findings were robust across sensitivity analyses using alternative costing methods, with only a few showing no differences between PRP and LFDF. These cost trends persisted in our exploratory analyses. A frequently cited concern is that orthobiologic treatments may not provide a definitive solution, but instead only postpone an inevitable surgical intervention. Rates of spine surgery following orthobiologic treatment were below the cell suppression threshold (>10 cases) at each follow-up, with our expanded analysis showing a rate of 3.8%. Even under conservative assumptions, these rates remain below or comparable to those observed among patients initially treated with surgery.^4,8,12^ While surgery remains essential for many patients with advanced degeneration, neurological deficits, or structural instability, these results indicate that expanding access to orthobiologic options when surgery is not the only viable choice could yield meaningful cost savings without increasing downstream utilization.

Pre-treatment differences in imaging and outpatient visits were notable, with higher utilization in the surgical cohort. This pattern could reflect greater condition severity in the surgical cohort but may also reflect more stringent diagnostic requirements for surgery, as imaging is often necessary for surgical planning and reimbursement, whereas it is not routinely required for orthobiologic treatment.

Consistent with this, surgical patients had higher rates of formal diagnoses supporting surgical indications (e.g., instability for fusion, stenosis for LFDF) and more frequent imaging in the 12 months prior to treatment. By most demographic, comorbidity, and diagnostic severity measures (e.g., multilevel involvement), the PRP cohort appeared comparable to the broader population undergoing fusion or LFDF, supporting generalizability of these findings. However, we cannot rule out the possibility that orthobiologic patients had a lower overall spinal condition burden compared to surgical patients. This limitation suggests that our findings may be most applicable to patients with lower symptom burden, a group for whom surgical intervention is often less clearly indicated.

We implemented rigorous matching procedures to create cohorts that were similar across demographic, health status, and prior health care use metrics. However, as with any non-randomized observational comparison, selection bias should be considered. Specifically, patients who choose and can afford orthobiologic treatment may differ in ways not captured by our data, and these unmeasured characteristics could make them more likely to respond favorably to conservative care.

Our intent was to develop two similar cohorts that would have been equally eligible for surgery or orthobiologic treatment. Matching achieved strong balance on characteristics likely related to clinical complexity but claims data cannot fully capture all factors influencing surgical decision-making. In particular, pain intensity, disability, chronicity, work status, and mental health can all influence treatment decision-making but these details were not available in our data. The absence of patient-reported outcomes limits our ability to make definitive conclusions about cost-effectiveness or value. Prior research indicates that patients often continue seeking care when initial treatments are ineffective.^21,22^ In our study, rates of subsequent treatment (i.e., re-injection in the PRP cohort and re-operation in the surgical cohort) were comparably low, suggesting that functional outcomes may have been similar across groups. While other studies support clinical improvements following PRP^25,26,31,32^, a direct comparison with surgical patients using patient-reported measures warrants further investigation.

The lack of high quality studies comparing surgery to PRP makes our findings challenging to fully interpret within the broader evidence base. A systematic review reported that ∼50–55% of patients achieve clinically meaningful pain relief at 6–12 months following orthobiologic treatment, but that review rates overall evidence quality as very low due to bias, small samples, and lack of blinding.^28^ Most studies have assessed adding orthobiologic treatment as an adjunct to surgery.^1,16,34^ A 2025 systematic review and meta-analysis of percutaneous endoscopic lumbar discectomy found that adding PRP to surgery reduced pain, disability, and recurrence rates compared with surgery alone.^34^ One small study (n=26) found rates of progression to spine surgery following orthobiologic treatment of 19% at 2 years and 23% at 3 years.^25,26^ Our observed rates were lower, although should be interpreted cautiously since we were only able to observe those who were continually enrolled in the health plan over the course of follow-up.

Several limitations should be considered when interpreting these findings. Generalizability may be affected by heterogeneity in orthobiologic preparation and the absence of standardized protocols, meaning results may not apply across all formulations or delivery methods. We combined laminectomy, foraminotomy, discectomy, and facetectomy into a single group due to similar indications and procedural characteristics; however, direct comparisons between orthobiologic treatment and individual procedures were not performed, partly because of limited sample sizes. Our analysis only included direct health care costs and focused on the payer perspective. Indirect costs, such as travel and lost work time, and out of pocket costs were not included; these costs would likely be higher for surgical patients given follow-up requirements and recovery periods.

Although the propensity-matched cohorts were balanced on measured characteristics, they represent a select subset of surgical candidates who elected orthobiologic treatment. Selection bias therefore remains possible, as patients choosing PRP may differ in expectations or other unmeasured factors associated with outcomes. In addition, the mean age of patients in our analytic sample (50–53 years) was younger than that reported in many population-based spine surgery studies, where average ages often exceed 60 years. We relied on proxy measures such as health care utilization intensity to approximate disease severity, as direct measures (e.g., imaging findings or patient-reported outcomes) were not available. While the matching process likely yielded cohorts with comparable clinical severity, the resulting sample reflects a younger population in whom the benefits of surgery for degenerative spine conditions may be less certain.

As with all claims-based analyses, assumptions were required regarding the extent to which billing data reflect underlying clinical decision-making. Interpretation was further limited by the fact that BMAC and PRP services were billed directly to Regenexx rather than captured through commercial payer claims, resulting in less granular diagnostic information for these patients. However, a standardized utilization review process was used to confirm that individuals met criteria for surgery, supporting comparability across cohorts. Finally, estimates at 36 and 48 months were based on smaller samples and should be interpreted cautiously. Larger, prospective studies using standardized protocols and incorporating patient-centered outcomes are needed to confirm these findings and inform coverage decisions.

In summary, our findings suggest that PRP may represent a cost-efficient alternative to spine surgery for select patients with degenerative spinal conditions, with meaningful savings and low rates of progression to surgery. Although surgery remains essential for some patients with advanced pathology, orthobiologic treatments may offer a lower-intensity option for appropriately selected individuals whose age or spine condition makes the benefit of surgery uncertain. Even modest reductions in surgical utilization could have important implications for health system and payer costs. These findings should be interpreted in light of key limitations, including potential selection bias, absence of patient-reported outcomes, and smaller samples at longer follow-up. Future research should focus on comparative effectiveness, long-term outcomes, and implementation strategies to support evidence-based coverage and reimbursement decisions.

## Data Availability

Data produced for the present study are not publicly available

## REFERENCES

1. Ajiboye RM, Hamamoto JT, Eckardt MA, Wang JC. Clinical and radiographic outcomes of concentrated bone marrow aspirate with allograft and demineralized bone matrix for posterolateral and interbody lumbar fusion in elderly patients. Eur Spine J. 2015;24(11):2567–2572. doi:10.1007/s00586-015-4117-5

2. Ambaliya S, De Bruyn F, Depreitere B. A systematic review and meta-analysis on surgery for lumbar disc herniation: optimal timing of surgery, return to work and outcomes compared with conservative management. Brain Spine. 2026;6:105917. doi:10.1016/j.bas.2025.105917

3. Belk JW, Lim JJ, Keeter C, et al. Patients With Knee Osteoarthritis Who Receive Platelet-Rich Plasma or Bone Marrow Aspirate Concentrate Injections Have Better Outcomes Than Patients Who Receive Hyaluronic Acid: Systematic Review and Meta-analysis. Arthroscopy. 2023;39(7):1714–1734. doi:10.1016/j.arthro.2023.03.001

4. Bullock G, Sangio CA, Beck EC, et al. Patient-Reported Outcomes and Reoperation Rates Following Lumbar Tubular Microdecompression: Six-year Follow-Up. Spine (Phila Pa 1976). 2023;48(5):350–357. doi:10.1097/BRS.0000000000004538

5. Carneiro D de C, Araújo LT de, Santos GC, et al. Clinical Trials with Mesenchymal Stem Cell Therapies for Osteoarthritis: Challenges in the Regeneration of Articular Cartilage. International Journal of Molecular Sciences. 2023;24(12). doi:10.3390/ijms24129939

6. Centeno C, Lucas M, Stemper I, Dodson E. Image Guided Injection of Anterior Cruciate Ligament Tears with Autologous Bone Marrow Concentrate and Platelets: Midterm Analysis from A Randomized Controlled Trial. Bio Orthop J. 2021;3(SP2):e7–e20. doi:10.22374/boj.v3iSP2.24

7. Centeno C, Sheinkop M, Dodson E, et al. A specific protocol of autologous bone marrow concentrate and platelet products versus exercise therapy for symptomatic knee osteoarthritis: a randomized controlled trial with 2 year follow-up. J Transl Med. 2018;16:355. doi:10.1186/s12967-018-1736-8

8. Cummins D, Hindoyan K, Wu HH, et al. Reoperation and Mortality Rates Following Elective 1 to 2 Level Lumbar Fusion: A Large State Database Analysis. Global Spine J. 2022;12(8):1708–1714. doi:10.1177/2192568220986148

9. Dieleman JL, Cao J, Chapin A, et al. US Health Care Spending by Payer and Health Condition, 1996-2016. JAMA. 2020;323(9):9. doi:10.1001/jama.2020.0734

10. Dos Santos RG, Santos GS, Alkass N, et al. The regenerative mechanisms of platelet-rich plasma: A review. Cytokine. 2021;144:155560. doi:10.1016/j.cyto.2021.155560

11. Dubin J, Leucht P, Murray M, Pezold R, Contributors S of the AA of OS on B of the PRP (PRP) for KOTOW and. American Academy of Orthopaedic Surgeons Technology Overview Summary: Platelet-Rich Plasma (PRP) for Knee Osteoarthritis. JAAOS - Journal of the American Academy of Orthopaedic Surgeons. 2024;32(7):296. doi:10.5435/JAAOS-D-23-00957

12. Etigunta SK, Liu AM, Gausper A, et al. Long-Term Reoperation Rates After Single-Level Lumbar Discectomy: A Nationwide Cohort Study. Spine (Phila Pa 1976). 2025;50(15):1052–1057. doi:10.1097/BRS.0000000000005328

13. Friedlis MF, Centeno CJ. Performing a Better Bone Marrow Aspiration. Phys Med Rehabil Clin N Am. 2016;27(4):919–939. doi:10.1016/j.pmr.2016.06.009

14. Goyal A, Payne S, Sangaralingham LR, et al. Incidence and risk factors for prolonged postoperative opioid use following lumbar spine surgery: a cohort study. Published online August 6, 2021. doi:10.3171/2021.2.SPINE202205

15. Hambright S, Koldewyn L, Donner C, Dregalla R, Herrera J, Donner J. Cellular And Acellular Characteristics Of Bone Marrow Aspirate Concentrate (Bmac) That Modulate Bone Marrow Derived Msc Health And Function. Osteoarthritis and Cartilage. 2023;31:S224–S225. doi:10.1016/j.joca.2023.01.211

16. Hart R, Komzák M, Okál F, Náhlík D, Jajtner P, Puskeiler M. Allograft alone versus allograft with bone marrow concentrate for the healing of the instrumented posterolateral lumbar fusion. The Spine Journal. 2014;14(7):1318–1324. doi:10.1016/j.spinee.2013.12.014

17. Hernigou P, Bouthors C, Bastard C, Flouzat Lachaniette CH, Rouard H, Dubory A. Subchondral bone or intra-articular injection of bone marrow concentrate mesenchymal stem cells in bilateral knee osteoarthritis: what better postpone knee arthroplasty at fifteen years? A randomized study. Int Orthop. 2021;45(2):391–399. doi:10.1007/s00264-020-04687-7

18. Hernigou P, Delambre J, Quiennec S, Poignard A. Human bone marrow mesenchymal stem cell injection in subchondral lesions of knee osteoarthritis: a prospective randomized study versus contralateral arthroplasty at a mean fifteen year follow-up. Int Orthop. 2021;45(2):365–373. doi:10.1007/s00264-020-04571-4

19. Kangari P, Talaei-Khozani T, Razeghian-Jahromi I, Razmkhah M. Mesenchymal stem cells: amazing remedies for bone and cartilage defects. Stem Cell Res Ther. 2020;11(1):492. doi:10.1186/s13287-020-02001-1

20. Kim GB, Seo MS, Park WT, Lee GW. Bone Marrow Aspirate Concentrate: Its Uses in Osteoarthritis. Int J Mol Sci. 2020;21(9):3224. doi:10.3390/ijms21093224

21. Lentz TA, Beneciuk JM, George SZ. Prediction of healthcare utilization following an episode of physical therapy for musculoskeletal pain. BMC Health Serv Res. 2018;18(1):1. doi:10.1186/s12913-018-3470-6

22. Mose S, Kent P, Smith A, Andersen JH, Christiansen DH. Trajectories of Musculoskeletal Healthcare Utilization of People with Chronic Musculoskeletal Pain &ndash; A Population-Based Cohort Study. CLEP. 2021;13:825–843. doi:10.2147/CLEP.S323903

23. Nawaz HMT, Jawwad MA, Khan MU, Qadir MJ, Farooq MO, Nawaz MH. Efficacy of Platelet-Rich Plasma Injections in Knee Osteoarthritis: A Systematic Review and Meta-Analysis. Cureus. 2025;17(10):e94288. doi:10.7759/cureus.94288

24. Nguyen AQ, Harvey JP, Khanna K, et al. Reasons for revision following stand-alone anterior lumbar interbody fusion and lateral lumbar interbody fusion. J Neurosurg Spine. 2021;35(1):60–66. doi:10.3171/2020.10.SPINE201239

25. Pettine K, Suzuki R, Sand T, Murphy M. Treatment of discogenic back pain with autologous bone marrow concentrate injection with minimum two year follow-up. Int Orthop. 2016;40(1):135–140. doi:10.1007/s00264-015-2886-4

26. Pettine KA, Suzuki RK, Sand TT, Murphy MB. Autologous bone marrow concentrate intradiscal injection for the treatment of degenerative disc disease with three-year follow-up. Int Orthop. 2017;41(10):2097–2103. doi:10.1007/s00264-017-3560-9

27. Philipp LR, Leibold A, Mahtabfar A, Montenegro TS, Gonzalez GA, Harrop JS. Achieving Value in Spine Surgery: 10 Major Cost Contributors. Global Spine J. 2021;11(1_suppl):14S–22S. doi:10.1177/2192568220971288

28. Schneider BJ, Hunt C, Conger A, et al. The effectiveness of intradiscal biologic treatments for discogenic low back pain: a systematic review. The Spine Journal. 2022;22(2):226–237. doi:10.1016/j.spinee.2021.07.015

29. Schug SA, Lavand’homme P, Barke A, et al. The IASP classification of chronic pain for ICD-11: chronic postsurgical or posttraumatic pain. Pain. 2019;160(1):1. doi:10.1097/j.pain.0000000000001413

30. Stark N, Kerr S, Stevens J. Prevalence and predictors of persistent post-surgical opioid use: a prospective observational cohort study. Anaesth Intensive Care. 2017;45(6):6. doi:10.1177/0310057X1704500609

31. Wang X, Wang S, Zhang J, Xie G, Zhang J. Platelet-rich plasma for the treatment of discogenic low back pain: a prospective randomized controlled trial. Front Pain Res. 2025;6. doi:10.3389/fpain.2025.1648772

32. Wolff M, Shillington JM, Rathbone C, Piasecki SK, Barnes B. Injections of concentrated bone marrow aspirate as treatment for Discogenic pain: a retrospective analysis. BMC Musculoskelet Disord. 2020;21(1):135. doi:10.1186/s12891-020-3126-7

33. Zaina F, Tomkins-Lane C, Carragee E, Negrini S. Surgical versus non-surgical treatment for lumbar spinal stenosis. Cochrane Database Syst Rev. 2016;2016(1):CD010264. doi:10.1002/14651858.CD010264.pub2

34. Zhang Y, Ju J, Wu J. Efficacy of platelet-rich plasma injection with percutaneous endoscopic lumbar discectomy for lumbar disc herniation: a systematic review and meta-analysis. Front Pharmacol. 2025;16. doi:10.3389/fphar.2025.1622974

35. Zhu X, Chan YT, Yung PSH, Tuan RS, Jiang Y. Subchondral Bone Remodeling: A Therapeutic Target for Osteoarthritis. Front Cell Dev Biol. 2021;8. doi:10.3389/fcell.2020.607764

