## Appendices for "Spine-Related Health Care Utilization and Costs Following Orthobiologic Injection Versus Lumbar Surgery for Degenerative Spine Conditions"

**Supplementary Appendix 1**

**Defining eligibility criterion for elective procedures preceded by a relevant clinical diagnosis**

A procedure was defined as elective and preceded by a relevant clinical diagnosis based on the following:

- Patient had at least one relevant diagnosis in an outpatient setting within 12 months preceding the Procedure. Relevant diagnoses are every M code listed under “Conditions of the lumbar spine compatible with DDD”
- If care was provided in the emergency department within 3 months prior to intervention, they must also have one relevant diagnosis in an outpatient setting occurring after the first service date of that ER visit and preceding the Procedure. If a patient had more than one emergency department visit in the prior 3 months, the visit closest to the procedure date must satisfy this requirement.
- If the procedure (surgery) occurs in the hospital (inpatient setting) it occurs on the first service date of the hospitalization. We would exclude inpatient index surgeries that occurred after the start date of the episode of care for the index surgery. The aim of this criterion is to exclude unplanned surgery or surgery that may occur incident to trauma.

**Procedure and diagnostic codes used for analysis**

**Table 1.** CPT codes for identifying spine procedures

| Procedure | CPT Code |
|---|---|
| Spinal Fusion | 22533, 22534, 22558, 22585, 22586, 22612, 22614, 22630, 22632, 22633, 22634, 63052, 63053 |
| Spinal fusion revision | 22849, 22850–22855 |
| Laminectomy, Foraminotomy, Discectomy, Facetectomy | 62380, 63005 and 63017, 63030, 63035, 63047, 63048 |
| Laminectomy, Foraminotomy, Discectomy, Facetectomy revision | 63042 |

**Table 2.** ICD-10 codes for lumbar DDD conditions

| Diagnosis | ICD-10-CM Code |
|---|---|
| Disc degeneration or spondylosis | M51.36, M51.37, M47.16, M47.17, M47.26, M47.27, M47.896, M47.897, M47.816, M47.817 |
| Spondylolithesis | M43.10, M43.16, M43.17, M43.19, M53.2_6, M53.2_7, M53.2_9 and "_" can be any value, M53.80, M53.86, M53.87 |
| Stenosis | M48.06, M48.07, M47.26, M47.27 |
| Myelopathy | M51.06, M51.07, M47.16, M47.17 |
| Radiculopathy | M54.16, M54.17, M47.26, M47.27, M51.16, M51.17, M54.3, M54.4 |

**Ineligible diagnoses (excluded)**

- Inflammatory syndrome diagnosis in the last 12 months (M04 – M14, M45.6, M45.7, M46). Examples of this group include
    - Inflammatory spondylopathies (M46)
    - Ankylosing spondylitis lumbar region (M45.6, M45.7)
    - Autoinflammatory syndromes (M04)
    - Rheumatoid arthritis (M05-M06)
    - Unspecified arthropathy (M12)
- Connective tissue disorders
    - Systemic connective tissue disorders (M30-M36)
    - Other disorders of the musculoskeletal system and connective tissue (M95)
- Infectious arthropathies (M00-M02)
- Injuries not suitable for orthobiologic procedures
    - Traumatic injuries (S12, S32–S34) to be included as a group, with specific examples below
        - Fractures (S32.0)
        - Dislocations (S33.1)
        - Lumbar spine fracture (S32)
        - Injury of spinal cord and nerves (S34)
        - Injury of Cervical vertebra by external causes (S12)
    - Other spondylopathies
        - Ankylosing hyperostosis (M48.1)
        - Traumatic spondylopathy (M48.3)
        - Fatigue fracture of vertebra (M48.4)
        - Collapsed vertebra (M48.5)
- Anterior spinal and vertebral artery compression syndromes (M47.0)
- Congenital, Neuromuscular and Idiopathic Spinal deformities
    - Idiopathic Scoliosis (M41.0, M41.1, M41.2)
    - Thoracogenic scoliosis (M41.3)
    - Neuromuscular Scoliosis (M41.4)
    - Congenital deformities including spine (Q67, Q76)
- Infections of the spine (e.g. osteomyelitis or discitis) (M46.2, M46.3, M46.4, M46.5, M46.8, M46.9) {Note: does not include M46.0 and M46.1 on purpose}
- Cancer: malignant neoplasms related to bone, spine or nervous system (C41, C47, C70, C71, C72, C79, C96)

**Definitions and identification of confounders used in analyses**

- Demographics
  - Geographic region
  - Age
  - Sex
- Conditions of the lumbar spine compatible with DDD {lumbar implied throughout below}
  - Disc degeneration or spondylosis. Combine any of the following:
    - Disc degeneration (M51.36, M51.37)
    - Spondylosis. Combine any of the following into spondylosis:
      - Spondylosis with myelopathy (M47.16, M47.17)
      - Spondylosis with radiculopathy (M47.26, M47.27)
      - Other Spondylosis (M47.896, M47.897)
      - Other spondylosis without myelopathy or radiculopathy (M47.816, M47.817)
  - Spondylolisthesis. Combine any of the following:
    - Spondylolisthesis (M43.10, M43.16, M43.17, M43.19*)*
    - Spinal instabilities (M53.2_6, M53.2_7, M53.2_9 and "_" can be any value)
    - Ligamentous instability (M53.80, M53.86, M53.87 M53.82)
  - Spine Stenosis (M48.06, M48.07) or foraminal narrowing (M47.26, M47.27)- *narrowing and leading to compression.* Combine any of the preceding into stenosis.
    - # of stenosis encounters in the prior 12 months
      - Count unique combination of (code, date)
  - Myelopathy. Combine:
    - Other disc disorders with myelopathy (M51.06, M51.07)
    - *Spondylosis* with myelopathy (M47.16, M47.17)
  - Radiculopathy. Combine:
    - Radiculopathy (M54.16, M54.17)
    - *Spondylosis* with radiculopathy (M47.26, M47.27)
    - Other disc disorders with Radiculopathy (M51.16, M51.17)
    - Sciatica (M54.3) or lumbago with sciatica (M54.4)
- Conditions of the lumbar spine not indicative of DDD if occuring in isolation (not with codes above)
  - Herniated disc / Intervertebral disc displacement (lumbar M51.26, lumbosacral M51.27)
  - Other lumbar/lumbosacral disc disorders (M51.86, M51.87)
  - Low back pain (dorsalgia of the lower back) (M54.5)
- General conditions of the spine (not specific to lumbar location)
  - Deforming dorsopathies (not idiopathic or congenital)
    - Spondylolysis (M43.0)
    - Secondary scoliosis (M41.5)
    - Other forms of scoliosis (M41.8)
    - Scoliosis unspecified (M41.9)
  - Kyphosis (M40)
- Multi-section conditions compatible with DDD {as surrogate for extent of DDD severity across spine}
  - Multi-section Disc degeneration (M51.3X with 1 X, 2 Xs; more than 2 Xs)
    - A categorical variable will be created with the following values:
      - No disc degeneration
      - Disc degeneration in one region
      - Disc degeneration in two regions
      - Disc degeneration in three or more regions
    - Similar variables will be created for other multi-section conditions
  - Multi-section Instability (M43.1X or M53.2_X with 1 X, 2 Xs; more than 2 Xs and "_" can be any value)
    - For example, a patient with M43.11 and M53.2_1 counts as having only one unique X because the X value for M43.11 is 1 and the X value for M53.2_1 is 1
    - M43.10 and M53.2_9 are both "site unspecified", so the 0 and the 9 do not count as two unique values in this context

        - M43.19 is "multiple sites in spine". Patients with M43.19 but less than two other unique Xs will be counted as having two unique Xs. Otherwise, the 9 in M43.19 will not count as a unique X value
        - Similar for similar variables
    - Multi-section Spinal stenosis (M48.0X with 1 X, 2 Xs; more than 2 Xs)
    - Multi-section Spondylosis (M47.1X or M47.2X or M47.89X or M47.81X with 1 X, 2 Xs; more than 2 Xs)
    - Multi-section radiculopathy (M54.1X or M47.2X or M51.1X with 1 X, 2 Xs; more than 2 Xs)
    - Multi-section myelopathy (M51.0X or M47.1X with 1 X, 2 Xs; more than 2 Xs)
- Musculoskeletal conditions (not exclusive to the spine) within 12 months prior to procedure
    - Poor bone quality. Combine:
        - Osteoporosis (M81)
        - Pathologic fracture (M80)
    - Osteoarthritis in other parts of the body. The following to be included as a single group.
        - Polyosteoarthritis (M15)
        - Osteoarthritis of the hip (M16)
        - Osteoarthritis knee (either place) (M17 codes)
        - Other osteoarthritis (M18 and M19)
- General comorbidity diagnosis in the 12 months prior to procedure
    - Obesity (E66.0, E66.1, E66.2, E66.8, E66.9, Z68.3, Z68.4, Z68.54, Z68.55, Z68.56)
    - Diabetes
        - Codes: E08 – E11, E13
    - Anxiety/Depression/Bipolar
        - Depression codes: F32, F33
        - Anxiety codes: F41
        - Bipolar codes: F31
- Medications within 12 months prior to the procedure
    - Prescription pain relievers included separately
        - Opioids
        - Prescription NSAIDs
        - Tramadol
        - Neuropathic pain meds (amitriptyline, duloxetine, pregabalin, gabapentin, venlafaxine)
- Intervention history in the 12 months prior to procedure (enumerated)
    - Diagnostic or therapeutic injection (CPT 62322 or 62323) [interlaminar injection into the space around the spinal nerves of lumbar or sacral region with/without imaging guidance]
    - Injection of an anesthetic agent or steroid (CPT 64483) [in the form of a transforaminal epidural into a single level of lumbar or sacral region]
        - Additional level Injection of an anesthetic agent or steroid (CPT 64484)
    - Use of oral corticosteroids
    - # of physical therapy encounters
    - Some less-invasive therapy: Any of the following (CPTs 62322, 62323, 64483, 64484, #PT encounters > 6)
- Health Seeking behavior / severity in the 12 months prior to procedure
    - # Outpatient medical visits in the past year
    - # ER visits in the prior year
    - # Chiropractic visits in the prior year
    - Spine xray
    - MRI of spine
- Other Severity proxies
    - Severity of stenosis – number of outpatient encounters with stenosis diagnosis code (M48.0) in the 12 months prior
    - Severity of disc degeneration - number of outpatient encounters with disc degeneration code (M51.3) in the 12 months prior
    - Severity of instability – number of outpatient encounters with instability (M43.1 or M53.2) in the 12 months prior

- Severity of osteoarthritis – number of outpatient encounters with OA diagnosis code of any joint in the 12 months prior to procedure (M15, M16, M17, M18, M19)
- Severity of orthopedic problems – number of outpatient encounters with any M diagnosis code in the in the 12 months prior to procedure

Descriptive data and absolute standardized mean differences (ASMDs) were reported on all the preceding variables. Given the limited sample size it was not possible to put all variables in the propensity score model. The following subset was used in propensity score matching because they were initially imbalanced (not balanced naturally) and not subject to reverse causation (not likely caused by preparation for surgery).

For the fusion comparison:

- Age
- Geographic region
- Sex
- Lumbar disc degeneration of spondylosis (Y/N)
- Lumbar spondylolisthesis (Y/N)
- Lumbar stenosis or foraminal narrowing (Y/N)
- Lumbar myelopathy (Y/N)
- Lumbar radiculopathy (Y/N)
- Lumbar herniated disc/intervertebral disc displacement (Y/N)
- Deforming dorsopathies (not specific to lumbar) (Y/N)
- Obesity (Y/N)
- Diabetes (Y/N)
- Depression/Anxiety/Bipolar (Y/N)
- Opioid or tramadol medication (Y/N)
- NSAID medication (Y/N)
- Neuropathic pain medication (Y/N)
- Diagnostic or therapeutic injection (CPT 62322, 62323) (Y/N)
- Injection of anesthetic or steroid (CPT 64483, 64484) (Y/N)
- Number of PT visits
- ER visit (Y/N)
- Chiropractic visit (Y/N)
- Number of instability outpatient visits
- Number of spinal stenosis outpatient visits
- Disc degeneration – number of regions
- Instability – number of regions
- Some less-invasive therapy (CPT 62322, 62323, 64483, 64484, GT 6 PT visits) (Y/N)

For the LFDF comparison:

- Age
- Geographic region
- Oral corticosteroids (Y/N)
- Lumbar disc degeneration or spondylosis (Y/N)
- Lumbar spondylolisthesis (Y/N)
- Lumbar stenosis or foraminal narrowing (Y/N)
- Lumbar myelopathy (Y/N)
- Lumbar radiculopathy (Y/N)
- Lumbar herniated disc/intervertebral disc displacement (Y/N)
- Deforming dorsopathies (Y/N)
- Obesity (Y/N)
- Diabetes (Y/N)
- Depression/Anxiety/Bipolar (Y/N)
- Opioid or tramadol medication (Y/N)
- NSAID medication (Y/N)
- Neuropathic pain medication (Y/N)
- Diagnostic or therapeutic injection (CPT 62322, 62323) (Y/N)

- Injection of anesthetic or steroid (CPT 64483, 64484) (Y/N)
- Number of PT visits
- ER visit (Y/N)
- Chiropractic visit (Y/N)
- Number of instability outpatient visits
- Number of spinal stenosis outpatient visits
- Disc degeneration – number of regions
- Instability – number of regions
- Some less-invasive therapy (CPT 62322, 62323, 64483, 64484, GT 6 PT visits) (Y/N)

**HCRU types and definitions**

We measured HCRU by interactions with the healthcare system (cumulative count data where appropriate). HCRU was captured longitudinally:

- From
  - The day after the Procedure END date
  - 90 days after the Procedure START date
    - Only to be used for bundled payment strategy when calculating costs for under DRG-based
- Until
  - 3 months after the Procedure START date
    - Only to be used for SNF/IRF/HH services that may be related to the procedure
  - 12 months after the Procedure START date
  - 24 months after the Procedure START date
  - 36 months after the Procedure START date
  - 48 months after the Procedure START date

The START and END dates of an index Procedure were defined as the START and END dates of that procedure's episode of care. Episodes of care were defined as follows:

- For outpatient claims, the episode of care starts and ends on the first service date of the claim
- For inpatient claims (including ER claims), claims that are not separated by at least one full day will count as part of the same episode of care. For example, see the table below

**Table 3.** Example of episode of care determination

| Patient | Claim place of service | Claim first service date | Claim last service date | Episode of care |
|---|---|---|---|---|
| A | ER | 1/1/2019 | 1/1/2019 | 1 |
| A | Inpatient | 1/2/2019 | 1/5/2019 | 1 |
| A | Inpatient | 1/4/2019 | 1/7/2019 | 1 |
| A | Inpatient | 1/9/2019 | 1/10/2019 | 2 |
| A | ER | 1/11/2019 | 1/12/2019 | 2 |
| A | Inpatient | 1/13/2019 | 1/14/2019 | 2 |

Some index procedures have both inpatient and outpatient claims. For example, a patient may have an outpatient claim with CPT code 22533 (spinal fusion) and an inpatient claim with CPT code 22534 (also spinal fusion) on the same day. In these cases, the procedure will be counted as inpatient.

Encounter types of interest are listed below. The number of encounters were counted as the number of unique episodes of care, except for radiology variables and subsequent BMAC and PRP procedures. For radiology variables, the number of encounters were counted as the number of unique claim First Service Dates. For subsequent BMAC and PRP procedures, the number of procedures were counted as the number of unique Primary Procedure Dates. Note that, for outpatient claims, counting unique episodes of care is the same thing as counting unique claim First Service Dates.

- Outpatient (non-physical therapy) visits with a relevant orthopedic diagnosis code
  - Relevant codes are the M and S ICD-10-CM codes listed in section 3.1 plus after care codes: Z96.651, Z96.652, Z47.1, Z47.89, Z48.89
  - Outpatient visits were considered non-physical therapy so long as a physical therapy outpatient visit did not occur on the same day
  - Note: because unique dates are used to distinguish a unique outpatient encounter multiple claims on the same day (e.g. professional and facility) were counted only once as a single encounter that day. All outpatient costs on that day were attributed to that encounter.
- Outpatient physical therapy with a relevant orthopedic diagnosis code

    - Relevant codes are the M and S ICD-10-CM codes plus after care codes: Z96.651, Z96.652, Z47.1, Z47.89, Z48.89
- Radiology regardless of contrast, i.e. with contrast and without contrast were grouped together
    - Xray of spine
    - MRI of spine
    - CT of spine
- Subsequent surgical spine procedures (occurrence of an eligible spine procedure during follow-up). Inclusive of codes for revision and draining. Outpatient procedures that occur during an inpatient episode of care will be counted as part of that episode of care for the purposes of counting unique encounters
    - Subsequent spinal fusion
    - Subsequent laminectomy, discectomy, foraminotomy, facetectomy
- Subsequent PRP and BMAC procedures
- Specialized care for wound healing. Outpatient claims that occur during an inpatient episode of care will be counted as part of that episode of care for the purposes of counting unique encounters. Combine:
    - Disruption of wound (T81.3)
    - Breakdown of sutures (T85.612)
    - Displacement of sutures (T85.622)
    - Procedural treatment for wound dehiscence (CPT 12020, 12021, 13160)
    - Specialized wound care (CPT 97597, 97598, 97602, 11042-11047)
- Emergency Department or Inpatient care for:
    - Post procedural complications
        - Pseudoarthrosis (M96.0)
        - Postlaminectomy syndrome (M96.1)
        - Other intraoperative and postprocedural complications (M96.8)
        - Unspecific complication of procedure (T81.8, T81.9)
        - Vascular complications following a procedure (T81.7)
    - Cerebrospinal fluid leaks
        - Cerebrospinal fluid leak (G96.0, G97.0) {may involve lumbar puncture as treatment CPT 62270 and 62272)}
        - Postprocedural cerebrospinal fluid leak (I97.83) {may involve lumbar puncture as treatment CPT 62270 and 62272)}
        - Cerebrospinal fluid leak detection and localization (CPT 78650)
        - Repair of dural/cerebrospinal fluid leak (63707, 63709)
    - Mechanical device complications (T84.226, T84.296) {from a cost perspective this will generate a revision}
    - Hematoma
    - Wound Infection
        - Infection following a procedure, inclusive of surgical site infection (T81.4)
        - Infections of the spine (e.g. osteomyelitis or discitis) (M46.2, M46.3, M46.4, M46.5, M46.8, M46.9)
        - Intracranial and intraspinal abscess or granuloma (G06)
        - Infection and inflammatory reaction due to internal fixation device of spine (T84.63)
    - Deep vein thrombosis, pulmonary embolism, or other post-surgical embolism
    - Sepsis
    - Inclusive of:
        - Number of stays
        - Number of inpatient days in hospital. Encounters that start and end on the same day will be considered to be one day long
- Skilled nursing facility services
    - POS code 31
- Inpatient rehabilitation facility services
    - POS code 61
- Home health services
    - POS code 12
- Opioid medications

- Number of unique prescription fill dates

### Costing

**Medicare cost methodology.** Costs are aggregated beginning at the PRPprocedure date and summed over time within the discrete time frame of interest. Costs will be assigned to each type of HCRU based on estimates of Medicare cost from external references and literature (Table 5). The Medicare cost of fusion/LFDF was generally reported as a 90-day bundled payment and therefore, no other costs were not counted until AFTER 90 days (that is no HCRU accrued a cost until after 90 days if fusion/LFDF was the initial procedure costed at $35,000/$9,000).

**Table 5. External cost assignment based on external literature**

| Service | Cost assigned | References |
|---|---|---|
| Fusion | $35,000 (90-day bundled) | [5, 7]$28,000-$46,000 based on DRG (450, 451, 457, 458)<br>[3]$30,000-$54,000 costs in Medicare population<br>[4]$45,000-$55,000 hospital costs |
| LFDF | $9,000 (90-day bundled) | [5]$6,000-$12,000 based on DRG (518, 519, 520)<br>[6]$12,000-$15,000 in-hospital cost for commercial payer<br>[6]$16,000-$24,000 over 90 days for commercial payer |
| Regenexx lumbar SCP | 2023: $7,636.75 | |
| Outpatient visit | $100 | [1]$89 - $128 office visit (99213), HOPD outpatient clinical visit (G0463) |
| Physical Therapy | $100 | [1]$31-34 per 15 minutes, assume 45 minute sessions |
| Back Xray | $50 | [1]$30-$70 (CPT 72100) |
| Back CT | $225 | [1]$150-$300 (CPT 72131,72132) |
| Back MRI | $240 | [2]$148-244 |
| Outpatient wound care | $150 | [1]$130-180 (CPT 97597, 11042) |
| Hospitalization DVT / PE / Embolism | $9,000 | [7]$9,000 (DRG 176) |
| Hospitalization Sepsis | $22,000 | [7]$11,000-$37,000 (DRG 870-871)<br>[8] $20,000 in 2018 plus approximately 25% for subsequent SNF care |
| Hospitalization for hematoma | $19,000 | [9]Not directly reported. Estimated as difference in Medicare payment for TKA with and without complication (17-22K) |
| Hospitalization for infection following a procedure | $22,000 | [7]$11,000-$37,000 (DRG 870-871)<br>[8] $20,000 in 2018 plus approximately 25% for subsequent SNF care |
| Hospitalization for post-procedural complications | $7,000 | [7]$4,000-$10,000 (DRG 919-921) |
| Hospitalization for cerebrospinal fluid leak | $7,000 | [7]$4,000-$10,000 (DRG 919-921) |
| Mechanical device complications | $7,000 | [7]$4,000-$10,000 (DRG 919-921) |

[1]CMS PFS Look-Up Tool https://www.cms.gov/medicare/physician-fee-schedule/search/overview
[2] https://www.medicare.gov/procedure-price-lookup/cost
[3]Malik AT, Phillips FM, Yu E, Khan SN. Are current DRG-based bundled payment models for lumbar fusions risk-adjusting adequately? An analysis of Medicare beneficiaries. The Spine Journal. 2020 Jan 1;20(1):32-40.

[4]Martin BI, Mirza SK, Karamian BA, Schoenfeld AJ, Ko H, Suri P, Brodke DS. Cost and Utilization Trends of Lumbar Fusion. JAMA Netw Open. 2026 Mar 2;9(3):e260452. doi: 10.1001/jamanetworkopen.2026.0452. PMID: 41779395; PMCID: PMC12961518.
[5]CMS FY 2025 files page / Table 5 and Table 1A-1E entry points (MS-DRG relative weights and standardized amounts used to verify the DRG planning anchors). https://www.cms.gov/medicare/payment/prospective-payment-systems/acute-inpatient-pps/acute-inpatient-files-download/files-fy-2025-final-rule-correction-notice
[6]Tiao, Justin BS[*]; Jain, Mayuri MPH[*,†]; Hoang, Ryan BS[*]; Yu, Alexander BS[*]; Huang, Jonathan J. AB[*]; Hecht, Andrew C. MD[*]; Stern, Brocha Z. PhD[*,†]; Chaudhary, Saad MD, MBA[*]. Immediate Procedure Reimbursement, 30-Day and 90-Day Episode Payments are Lower in Ambulatory Surgery Centers Versus Hospital Outpatient Departments for Lumbar Laminectomy. Clinical Spine Surgery
[7]https://www.optumcoding.com/upload/docs/2021%20DRG_National%20Average%20Payment%20Table_Update.pdf?srsltid=AfmBOorWJgoPa0W1CBB70W3N3alJOvSXoDnJ1r_PC2pgLwNGmjMwAhiK&utm_source=chatgpt.com
[8]Buchman TG, Simpson SQ, Sciarretta KL, Finne KP, Sowers N, Collier M, Chavan S, Oke I, Pennini ME, Santhosh A, Wax M. Sepsis among medicare beneficiaries: 1. The burdens of sepsis, 2012–2018. Critical care medicine. 2020 Mar 1;48(3):276-88.
[9]Haidar S, Vazquez R, Medic G. Impact of surgical complications on hospital costs and revenues: retrospective database study of Medicare claims. Journal of comparative effectiveness research. 2023 Jul;12(7):e230080.

**Multiplier methodology.** To illustrate a potential private payer cost, Medicare based costs were multiplied by 2 (which is in the range of multipliers across different cost settings).

**Table 6. Multipliers used for private payer costing estimate**

| Service Type | Commercial:Medicare Payment Ratio | Source (Typical) |
|---|---|---|
| Inpatient hospital | 1.8 to 2.5 | RAND Hospital Price Transparency Studies |
| Outpatient hospital | 2.0 to 3.0 | MedPAC, RAND, HCCI |
| Physician services | 1.2 to 1.8 | MedPAC, CMS Office of the Actuary |
| Ambulatory surgery | 1.4 to 2.0 | HCCI, FAIR Health |

**Private payer average cost methodology.** Using COMMERCIAL PAYER data, we obtained the average cost for an episode of care associated with each type of healthcare resource. For example, a surgical procedure might have physician billing and facility billing separately, along with a different bill for anesthesia as for the surgeon. Multiple claims for the same episode of care will be treated as a single health care use (and counted towards more expensive/acute visit type) and all applied costs will be summed to compute the total cost of care. Table 7 shows the average cost of care for each type of service among patients receiving orthopedic care in the COMMERCIAL PAYER data.

**Table 7. Commercial payer cost assignment; based average allowed amounts in commercial data**

| Service | Cost assigned |
|---|---|
| Fusion | $48,159 |
| LFDF | $20,580 |
| Regenexx lumbar PRP | 2023: $7,636.75 |
| Outpatient visit | $444 |
| Physical Therapy | $107 |
| Spine xray | $103 |
| Spine CT | $530 |
| Spine MRI | $822 |
| Outpatient wound care | $1,346 |
| Hospitalization DVT / PE / Embolism | $35,308 |

| | |
|---|---|
| Hospitalization Sepsis | $44,514 |
| Hospitalization for hematoma | $39,130 |
| Hospitalization for infection following a procedure | $43,976 |
| Hospitalization for post-procedural complications | $32,228 |
| Hospitalization for cerebrospinal fluid leak | $39,696 |
| Mechanical device complications | $34,456 |

**Computation of aggregate costs.**

Each occurrence of a relevant HCRU was assigned a cost and all costs were aggregated over the discrete time frame of interest (1,2,3,4 years). The cost of the initial procedure was also estimated and included in the sum.  When calculating Medicare costs, reimbursement for inpatient procedures is typically based on a bundled payment program (lump sum that covers all related costs for 90 days) rather than fee for service (or individual service costs). This impacted the published estimates of costs for surgery are frequently an inpatient service. The literature on surgery reports a bundled payment. Therefore, we used that bundled payment amount for the cost of surgery (in the Medicare framework) and did not accrue any additional costs until after 90 days, when costing surgery. Overlapping records of identical visit types were treated as a single health care use (and counted towards more expensive/acute visit type) and all applied costs were summed; e.g. two outpatient visits on same day counted once towards the tally of outpatient visit HCRU type, and the associated cost of this single tally was the sum of any costs at either visit. A lack of utilization data was treated as zero (0) utilization and thus attributed $0 in costs.

**Supplementary Table 1. Selection of analytic sample**

| Inclusion/exclusion criteria | Orthobiologic Procedures | Fusion Procedures | LFDF Procedures |
|---|---|---|---|
| Starting population: Receiving eligible procedures | 3470 | 184,721 | 235,989 |
| Procedure occurred between 1/1/2016 and 12/31/2023 | 3217 | 177,371 | 228,356 |
| Exclude patients who have multiple DOBs or sexes in the data | 3216 | 177,368 | 228,349 |
| Patients age 18 or older at the time of the procedure | 3205 | 176,328 | 227,538 |
| 12 months of continuous enrollment with commercial insurer both before and after the procedure | 824 | 85,826 | 115,219 |
| At least one outpatient claim in the 12 months prior to the procedure | 802 | 85,796 | 115,211 |
| At least one outpatient claim in the 12 months after the procedure | 783 | 85,750 | 115,154 |
| **Procedure is elective and preceded by a relevant clinical diagnosis** | | | |
| At least one relevant diagnosis (DXs compatible with DDD) in the 12 months prior to the procedure | 593 | 76,177 | 112,113 |
| If ER care in the 3 months prior to the procedure, at least one relevant DDD dx in an outpatient setting after the ER care but prior to the procedure | 582 | 72,915 | 106,547 |
| Filter out emergent procedures for complications arising in the hospital | 582 | 66,055 | 104,557 |
| **Exclude patients with clinical contra-indications** | | | |
| Inflammatory syndrome dx in the 12 months prior to the procedure | 433 | 45,967 | 82,531 |
| Ineligible connective tissue disorder dx in the 12 months prior to the procedure | 416 | 45,014 | 81,385 |
| Infectious arthropathy dx in the 12 months prior to the procedure | 416 | 44,950 | 81,331 |
| Traumatic injury not suitable for Regenexx dx in the 12 months prior to the procedure | 370 | 41,274 | 75,730 |
| Other spondylopathies dx in the 12 months prior to the procedure | 366 | 40,242 | 74,957 |
| Anterior spinal or vertebral artery compression syndrome dx in the 12 months prior to the procedure | 366 | 40,184 | 74,929 |
| Congenital, neuromuscular, or idiopathic spinal deformity dx in the 12 months prior to the procedure | 349 | 36,977 | 72,685 |
| Spine infection dx in the 12 months prior to the procedure | 349 | 36,977 | 72,685 |
| Cancer: malignant neoplasms related to bone, spine, or nervous system | 349 | 36,843 | 72,483 |
| **Exclude patients with prior treatment contra-indications** | | | |
| Exclude Regenexx patients who had corticosteroids in the prior 2 months | 310 | 36,843 | 72,483 |
| Previous spine surgery in the 12 months prior to the procedure | 307 | 33,133 | 60,968 |
| RFA of spine overall in the 12 months prior to the procedure | 298 | 31,859 | 60,026 |
| Intraoperative and post-procedural complications and disorders of the musculoskeletal system in the 12 months prior to the procedure | 287 | 27,131 | 57,761 |
| Received mab medication in the 3 months prior to the procedure | 285 | 26,665 | 57,112 |
| Exclude surgery patients who had Regenexx at any point | 285 | 26,652 | 57,074 |
| If a patient had multiple procedures, take the first one | 262 | 25,778 | 56,374 |

DOB, date of birth; Dx, diagnosis; DDD, degenerative disc disease; ER, emergency room; RFA, radio frequency ablation

**Supplementary Table 2. Pre-matching demographic and clinical characteristics – PRPversus surgery**

| **Characteristic** | **PRP**<br>N = 262 | **Spine Surgery**<br>N = 82,152 |
|---|---|---|
| Treatment | | |
| BMAC/PRP | 262 (100.0%) | -- |
| Spinal fusion | -- | 25,778 (31.4%) |
| Laminectomy, foraminotomy, discectomy, facetectomy | -- | 56,374 (68.6%) |
| Continuous commercial health plan enrollment after index procedure | | |
| CE for 12 months | 262 (100.0%) | 82,152 (100.0%) |
| CE for 24 months | 165 (63.0%) | 56,671 (69.0%) |
| CE for 36 months | 110 (42.0%) | 38,510 (46.9%) |
| CE for 48 months | 70 (26.7%) | 24,956 (30.4%) |
| Sex | | |
| Female | 114 (43.5%) | 35,541 (43.3%) |
| Male | 148 (56.5%) | 46,611 (56.7%) |
| Age | | |
| Mean (SD) | 50.5 (11.6) | 50.8 (11.4) |
| Median (Q1, Q3) | 53.0 (43.0, 60.0) | 53.0 (43.0, 60.0) |
| Geographic region | | |
| Northeast | 59 (22.5%) | 14,805 (18.0%) |
| Midwest | 94 (35.9%) | 36,561 (44.5%) |
| South | 60 (22.9%) | 23,519 (28.6%) |
| West | 49 (18.7%) | 7,212 (8.8%) |
| Other | <11 | 55 (0.1%) |
| Conditions of the lumbar spine compatible with DDD within 12 months prior to the procedure | | |
| Lumbar disc degeneration or spondylosis | 209 (79.8%) | 62,675 (76.3%) |
| Lumbar spondylolisthesis | 43 (16.4%) | 23,898 (29.1%) |
| Lumbar stenosis or foraminal narrowing | 99 (37.8%) | 58,454 (71.2%) |
| Lumbar myelopathy | <11 | 3,207 (3.9%) |
| Lumbar radiculopathy | 189 (72.1%) | 75,812 (92.3%) |
| Conditions of the lumbar spine not indicative of DDD within 12 months prior to the procedure | | |
| Herniated disc/intervertebral disc displacement | 100 (38.2%) | 52,805 (64.3%) |
| Other lumbar/lumbosacral disc disorders | <11 | 2,414 (2.9%) |
| Low back pain | 178 (67.9%) | 62,674 (76.3%) |
| Conditions related to the spine (not specific to lumbar) within 12 months prior to the procedure | | |
| Deforming dorsopathies | 12 (4.6%) | 7,566 (9.2%) |
| Kyphosis | <11 | 940 (1.1%) |
| Multi-section conditions compatible with DDD within 12 months prior to the procedure | | |
| Disc degeneration | 138 (52.7%) | 44,998 (54.8%) |
| Disc degeneration - number of regions | | |
| One region | 120 (45.8%) | 34,706 (42.2%) |
| Two regions | 18 (6.9%) | 9,939 (12.1%) |
| Three or more regions | <11 | 353 (0.4%) |
| None | 124 (47.3%) | 37,154 (45.2%) |
| Instability | 40 (15.3%) | 23,108 (28.1%) |
| Instability - number of regions | | |
| One region | 32 (12.2%) | 16,985 (20.7%) |
| Two regions | <11 | 5,341 (6.5%) |
| Three or more regions | <11 | 782 (1.0%) |

| | | |
|---|---|---|
| None | 222 (84.7%) | 59,044 (71.9%) |
| Spinal stenosis | 83 (31.7%) | 55,578 (67.7%) |
| Spinal stenosis - number of regions | | |
| One region | 70 (26.7%) | 41,400 (50.4%) |
| Two regions | 12 (4.6%) | 12,409 (15.1%) |
| Three or more regions | <11 | 1,769 (2.2%) |
| None | 179 (68.3%) | 26,574 (32.3%) |
| Spondylosis | 154 (58.8%) | 48,590 (59.1%) |
| Spondylosis - number of regions | | |
| One region | 106 (40.5%) | 33,101 (40.3%) |
| Two regions | 45 (17.2%) | 13,156 (16.0%) |
| Three or more regions | <11 | 2,333 (2.8%) |
| None | 108 (41.2%) | 33,562 (40.9%) |
| Radiculopathy | 184 (70.2%) | 73,803 (89.8%) |
| Radiculopathy - number of regions | | |
| One region | 123 (46.9%) | 39,552 (48.1%) |
| Two regions | 48 (18.3%) | 28,838 (35.1%) |
| Three or more regions | 13 (5.0%) | 5,413 (6.6%) |
| None | 78 (29.8%) | 8,349 (10.2%) |
| Myelopathy | <11 | 4,826 (5.9%) |
| Myelopathy - number of regions | | |
| One region | <11 | 4,621 (5.6%) |
| Two regions | <11 | 188 (0.2%) |
| Three or more regions | <11 | 17 (0.0%) |
| None | 258 (98.5%) | 77,326 (94.1%) |
| Musculoskeletal conditions (not exclusive to the spine) within 12 months prior to the procedure | | |
| Poor bone quality | <11 | 1,369 (1.7%) |
| Osteoarthritis in other parts of the body | 66 (25.2%) | 15,286 (18.6%) |
| General comorbidity diagnoses within 12 months prior to the procedure | | |
| Obesity | 30 (11.5%) | 22,353 (27.2%) |
| Diabetes | 16 (6.1%) | 12,638 (15.4%) |
| Depression/Anxiety/Bipolar | 53 (20.2%) | 20,805 (25.3%) |
| Medications within 12 months prior to the procedure | | |
| Opioid or tramadol medication | 73 (27.9%) | 32,637 (39.7%) |
| NSAID medication | 57 (21.8%) | 29,136 (35.5%) |
| Neuropathic pain medication | 37 (14.1%) | 25,274 (30.8%) |
| Intervention history in the 12 months prior to the procedure | | |
| Diagnostic or therapeutic injection (CPT 62322, 62323) | 14 (5.3%) | 16,301 (19.8%) |
| Injection or anesthetic or steroid (CPT 64483, 64484) | 33 (12.6%) | 26,457 (32.2%) |
| Oral corticosteroids | 59 (22.5%) | 34,131 (41.5%) |
| Physical therapy | 147 (56.1%) | 43,208 (52.6%) |
| Greater than 6 PT visits | 84 (32.1%) | 20,887 (25.4%) |
| Number of PT visits | | |
| Mean (SD) | 6.8 (10.4) | 5.0 (8.9) |
| Median (Q1, Q3) | 1.0 (0.0, 10.0) | 1.0 (0.0, 7.0) |
| Health seeking behavior in the 12 months prior to the procedure | | |
| Number of outpatient visits | | |
| Mean (SD) | 22.6 (15.6) | 23.6 (15.6) |
| Median (Q1, Q3) | 19.0 (11.0, 31.0) | 20.0 (13.0, 30.0) |
| ER visit | 36 (13.7%) | 15,755 (19.2%) |
| Number of ER visits | | |

| | | |
|---|---|---|
| Mean (SD) | 0.2 (0.5) | 0.3 (0.8) |
| Median (Q1, Q3) | 0.0 (0.0, 0.0) | 0.0 (0.0, 0.0) |
| Chiropractic visit | 79 (30.2%) | 19,810 (24.1%) |
| Number of chiropractic visit | | |
| Mean (SD) | 3.6 (8.1) | 2.2 (6.0) |
| Median (Q1, Q3) | 0.0 (0.0, 2.0) | 0.0 (0.0, 0.0) |
| Spine x-ray | 123 (46.9%) | 61,615 (75.0%) |
| Spine MRI | 178 (67.9%) | 75,916 (92.4%) |
| Spine x-ray or spine MRI | 204 (77.9%) | 79,345 (96.6%) |
| Other severity proxies in the 12 months prior to the procedure | | |
| Spinal stenosis outpatient visit | 83 (31.7%) | 55,406 (67.4%) |
| Number of spinal stenosis outpatient visits | | |
| Mean (SD) | 0.7 (1.7) | 2.1 (3.1) |
| Median (Q1, Q3) | 0.0 (0.0, 1.0) | 1.0 (0.0, 3.0) |
| Disc degeneration outpatient visit | 138 (52.7%) | 44,878 (54.6%) |
| Number of disc degeneration outpatient visits | | |
| Mean (SD) | 1.6 (3.7) | 1.6 (3.2) |
| Median (Q1, Q3) | 1.0 (0.0, 2.0) | 1.0 (0.0, 2.0) |
| Instability outpatient visit | 40 (15.3%) | 23,036 (28.0%) |
| Number of instability outpatient visits | | |
| Mean (SD) | 0.4 (1.7) | 0.9 (2.4) |
| Median (Q1, Q3) | 0.0 (0.0, 0.0) | 0.0 (0.0, 1.0) |
| Osteoarthritis outpatient visit | 66 (25.2%) | 15,137 (18.4%) |
| Number of osteoarthritis outpatient visits | | |
| Mean (SD) | 0.9 (3.1) | 0.5 (1.9) |
| Median (Q1, Q3) | 0.0 (0.0, 1.0) | 0.0 (0.0, 0.0) |
| Less invasive intervention indicator (CPT 62322, 62323, 64483, 64484, GT 6 PT visits) | 115 (43.9%) | 47,658 (58.0%) |

CE, continuous enrollment; DDD, degenerative disc disease; SD, standard deviation; CPT, current procedural terminology; MRI, magnetic resonance imaging; PT, physical therapy

**Supplementary Table 3. Pre-matching demographic and clinical characteristics – PRP versus fusion vs LFDF**

| **Characteristic** | **PRP** N = 262 | **Fusion** N = 25,778 | **LFDF** N = 56,374 |
|---|---|---|---|
| Continuous commercial health plan enrollment after index procedure | | | |
| CE for 12 months | 262 (100.0%) | 25,778 (100.0%) | 56,374 (100.0%) |
| CE for 24 months | 165 (63.0%) | 16,962 (65.8%) | 39,709 (70.4%) |
| CE for 36 months | 110 (42.0%) | 11,113 (43.1%) | 27,397 (48.6%) |
| CE for 48 months | 70 (26.7%) | 6,907 (26.8%) | 18,049 (32.0%) |
| Sex | | | |
| Female | 114 (43.5%) | 13,341 (51.8%) | 22,200 (39.4%) |
| Male | 148 (56.5%) | 12,437 (48.2%) | 34,174 (60.6%) |
| Age | | | |
| Mean (SD) | 50.5 (11.6) | 54.5 (9.3) | 49.1 (11.8) |
| Median (Q1, Q3) | 53.0 (43.0, 60.0) | 56.0 (49.0, 61.0) | 51.0 (41.0, 59.0) |
| Geographic region | | | |
| Northeast | 59 (22.5%) | 4,541 (17.6%) | 10,264 (18.2%) |
| Midwest | 94 (35.9%) | 11,509 (44.6%) | 25,052 (44.4%) |
| South | 60 (22.9%) | 7,586 (29.4%) | 15,933 (28.3%) |
| West | 49 (18.7%) | 2,127 (8.3%) | 5,085 (9.0%) |
| Other | <11 | 15 (0.1%) | 40 (0.1%) |
| Conditions of the lumbar spine compatible with DDD within 12 months prior to the procedure | | | |
| Lumbar disc degeneration or spondylosis | 209 (79.8%) | 21,517 (83.5%) | 41,158 (73.0%) |
| Lumbar spondylolisthesis | 43 (16.4%) | 15,720 (61.0%) | 8,178 (14.5%) |
| Lumbar stenosis or foraminal narrowing | 99 (37.8%) | 20,779 (80.6%) | 37,675 (66.8%) |
| Lumbar myelopathy | <11 | 960 (3.7%) | 2,247 (4.0%) |
| Lumbar radiculopathy | 189 (72.1%) | 22,648 (87.9%) | 53,164 (94.3%) |
| Conditions of the lumbar spine not indicative of DDD within 12 months prior to the procedure | | | |
| Herniated disc/intervertebral disc displacement | 100 (38.2%) | 12,509 (48.5%) | 40,296 (71.5%) |
| Other lumbar/lumbosacral disc disorders | <11 | 744 (2.9%) | 1,670 (3.0%) |
| Low back pain | 178 (67.9%) | 19,770 (76.7%) | 42,904 (76.1%) |
| Conditions related to the spine (not specific to lumbar) within 12 months prior to the procedure | | | |
| Deforming dorsopathies | 12 (4.6%) | 4,187 (16.2%) | 3,379 (6.0%) |
| Kyphosis | <11 | 477 (1.9%) | 463 (0.8%) |
| Multi-section conditions compatible with DDD within 12 months prior to the procedure | | | |
| Disc degeneration | 138 (52.7%) | 15,931 (61.8%) | 29,067 (51.6%) |
| Disc degeneration - number of regions | | | |
| One region | 120 (45.8%) | 11,855 (46.0%) | 22,851 (40.5%) |
| Two regions | 18 (6.9%) | 3,923 (15.2%) | 6,016 (10.7%) |
| Three or more regions | <11 | 153 (0.6%) | 200 (0.4%) |
| None | 124 (47.3%) | 9,847 (38.2%) | 27,307 (48.4%) |
| Instability | 40 (15.3%) | 15,760 (61.1%) | 7,348 (13.0%) |
| Instability - number of regions | | | |
| One region | 32 (12.2%) | 10,741 (41.7%) | 6,244 (11.1%) |
| Two regions | <11 | 4,323 (16.8%) | 1,018 (1.8%) |
| Three or more regions | <11 | 696 (2.7%) | 86 (0.2%) |
| None | 222 (84.7%) | 10,018 (38.9%) | 49,026 (87.0%) |

| | | | |
|---|---|---|---|
| Spinal stenosis | 83 (31.7%) | 20,634 (80.0%) | 34,944 (62.0%) |
| Spinal stenosis - number of regions | | | |
| One region | 70 (26.7%) | 14,298 (55.5%) | 27,102 (48.1%) |
| Two regions | 12 (4.6%) | 5,420 (21.0%) | 6,989 (12.4%) |
| Three or more regions | <11 | 916 (3.6%) | 853 (1.5%) |
| None | 179 (68.3%) | 5,144 (20.0%) | 21,430 (38.0%) |
| Spondylosis | 154 (58.8%) | 18,030 (69.9%) | 30,560 (54.2%) |
| Spondylosis - number of regions | | | |
| One region | 106 (40.5%) | 11,236 (43.6%) | 21,865 (38.8%) |
| Two regions | 45 (17.2%) | 5,588 (21.7%) | 7,568 (13.4%) |
| Three or more regions | <11 | 1,206 (4.7%) | 1,127 (2.0%) |
| None | 108 (41.2%) | 7,748 (30.1%) | 25,814 (45.8%) |
| Radiculopathy | 184 (70.2%) | 22,261 (86.4%) | 51,542 (91.4%) |
| Radiculopathy - number of regions | | | |
| One region | 123 (46.9%) | 12,346 (47.9%) | 27,206 (48.3%) |
| Two regions | 48 (18.3%) | 8,141 (31.6%) | 20,697 (36.7%) |
| Three or more regions | 13 (5.0%) | 1,774 (6.9%) | 3,639 (6.5%) |
| None | 78 (29.8%) | 3,517 (13.6%) | 4,832 (8.6%) |
| Myelopathy | <11 | 1,945 (7.5%) | 2,881 (5.1%) |
| Myelopathy - number of regions | | | |
| One region | <11 | 1,830 (7.1%) | 2,791 (5.0%) |
| Two regions | <11 | 104 (0.4%) | 84 (0.1%) |
| Three or more regions | <11 | 11 (0.0%) | <11 |
| None | 258 (98.5%) | 23,833 (92.5%) | 53,493 (94.9%) |
| Musculoskeletal conditions (not exclusive to the spine) within 12 months prior to the procedure | | | |
| Poor bone quality | <11 | 751 (2.9%) | 618 (1.1%) |
| Osteoarthritis in other parts of the body | 66 (25.2%) | 6,313 (24.5%) | 8,973 (15.9%) |
| General comorbidity diagnoses within 12 months prior to the procedure | | | |
| Obesity | 30 (11.5%) | 8,064 (31.3%) | 14,289 (25.3%) |
| Diabetes | 16 (6.1%) | 4,870 (18.9%) | 7,768 (13.8%) |
| Depression/Anxiety/Bipolar | 53 (20.2%) | 7,627 (29.6%) | 13,178 (23.4%) |
| Medications within 12 months prior to the procedure | | | |
| Opioid or tramadol medication | 73 (27.9%) | 9,776 (37.9%) | 22,861 (40.6%) |
| NSAID medication | 57 (21.8%) | 8,781 (34.1%) | 20,355 (36.1%) |
| Neuropathic pain medication | 37 (14.1%) | 8,028 (31.1%) | 17,246 (30.6%) |
| Intervention history in the 12 months prior to the procedure | | | |
| Diagnostic or therapeutic injection (CPT 62322, 62323) | 14 (5.3%) | 5,100 (19.8%) | 11,201 (19.9%) |
| Injection or anesthetic or steroid (CPT 64483, 64484) | 33 (12.6%) | 7,688 (29.8%) | 18,769 (33.3%) |
| Oral corticosteroids | 59 (22.5%) | 9,005 (34.9%) | 25,126 (44.6%) |
| Physical therapy | 147 (56.1%) | 12,669 (49.1%) | 30,539 (54.2%) |
| Greater than 6 PT visits | 84 (32.1%) | 6,126 (23.8%) | 14,761 (26.2%) |
| Number of PT visits | | | |
| Mean (SD) | 6.8 (10.4) | 4.7 (8.8) | 5.2 (8.9) |
| Median (Q1, Q3) | 1.0 (0.0, 10.0) | 0.0 (0.0, 6.0) | 1.0 (0.0, 7.0) |
| Less invasive intervention indicator (CPT 62322, 62323, 64483, 64484, GT 6 PT visits) | 115 (43.9%) | 14,412 (55.9%) | 33,246 (59.0%) |
| Health seeking behavior in the 12 months prior to the procedure | | | |
| Number of outpatient visits | | | |

| | | | |
|---|---|---|---|
| Mean (SD) | 22.6 (15.6) | 25.6 (16.0) | 22.6 (15.3) |
| Median (Q1, Q3) | 19.0 (11.0, 31.0) | 22.0 (15.0, 32.0) | 19.0 (12.0, 29.0) |
| ER visit | 36 (13.7%) | 4,498 (17.4%) | 11,257 (20.0%) |
| Number of ER visits | | | |
| Mean (SD) | 0.2 (0.5) | 0.3 (0.8) | 0.3 (0.8) |
| Median (Q1, Q3) | 0.0 (0.0, 0.0) | 0.0 (0.0, 0.0) | 0.0 (0.0, 0.0) |
| Chiropractic visit | 79 (30.2%) | 4,629 (18.0%) | 15,181 (26.9%) |
| Number of chiropractic visit | | | |
| Mean (SD) | 3.6 (8.1) | 1.7 (5.4) | 2.4 (6.3) |
| Median (Q1, Q3) | 0.0 (0.0, 2.0) | 0.0 (0.0, 0.0) | 0.0 (0.0, 1.0) |
| Spine x-ray | 123 (46.9%) | 21,157 (82.1%) | 40,458 (71.8%) |
| Spine MRI | 178 (67.9%) | 23,374 (90.7%) | 52,542 (93.2%) |
| Spine x-ray or spine MRI | 204 (77.9%) | 24,882 (96.5%) | 54,463 (96.6%) |
| Other severity proxies in the 12 months prior to the procedure | | | |
| Spinal stenosis outpatient visit | 83 (31.7%) | 20,557 (79.7%) | 34,849 (61.8%) |
| Number of spinal stenosis outpatient visits | | | |
| Mean (SD) | 0.7 (1.7) | 3.0 (3.6) | 1.8 (2.7) |
| Median (Q1, Q3) | 0.0 (0.0, 1.0) | 2.0 (1.0, 4.0) | 1.0 (0.0, 2.0) |
| Disc degeneration outpatient visit | 138 (52.7%) | 15,885 (61.6%) | 28,993 (51.4%) |
| Number of disc degeneration outpatient visits | | | |
| Mean (SD) | 1.6 (3.7) | 2.0 (3.5) | 1.5 (3.1) |
| Median (Q1, Q3) | 1.0 (0.0, 2.0) | 1.0 (0.0, 2.0) | 1.0 (0.0, 2.0) |
| Instability outpatient visit | 40 (15.3%) | 15,708 (60.9%) | 7,328 (13.0%) |
| Number of instability outpatient visits | | | |
| Mean (SD) | 0.4 (1.7) | 2.2 (3.3) | 0.3 (1.4) |
| Median (Q1, Q3) | 0.0 (0.0, 0.0) | 1.0 (0.0, 3.0) | 0.0 (0.0, 0.0) |
| Osteoarthritis outpatient visit | 66 (25.2%) | 6,237 (24.2%) | 8,900 (15.8%) |
| Number of osteoarthritis outpatient visits | | | |
| Mean (SD) | 0.9 (3.1) | 0.7 (2.4) | 0.4 (1.7) |
| Median (Q1, Q3) | 0.0 (0.0, 1.0) | 0.0 (0.0, 0.0) | 0.0 (0.0, 0.0) |
| Less invasive intervention indicator (CPT 62322, 62323, 64483, 64484, GT 6 PT visits) | 115 (43.9%) | 14,412 (55.9%) | 33,246 (59.0%) |

CE, continuous enrollment; DDD, degenerative disc disease; SD, standard deviation; CPT, current procedural terminology; MRI, magnetic resonance imaging; PT, physical therapy

**Supplementary Table 4. Healthcare resource utilization 36 months post procedure – PRP versus fusion**

| **Characteristic** | **PRP**<br>N = 41 | **Fusion**<br>N = 1,150 | **p-value** |
|---|---|---|---|
| Number of outpatient visits | | | 0.11 |
| Mean (SD) | 10.4 (12.4) | 11.8 (12.0) | |
| Median (Q1, Q3) | 5.0 (3.0, 15.0) | 8.0 (4.0, 16.0) | |
| Number of outpatient physical therapy visits | | | 0.96 |
| Mean (SD) | 10.9 (24.7) | 8.4 (16.3) | |
| Median (Q1, Q3) | 1.0 (0.0, 6.0) | 2.0 (0.0, 11.0) | |
| Spine x-ray | 16 (39.0%) | 1,104 (96.0%) | < 0.001 |
| MRI of spine | 14 (34.1%) | 481 (41.8%) | 0.41 |
| CT of spine | <11 | 310 (27.0%) | 0.05 |
| Subsequent fusion | <11 | 135 (11.7%) | -- |
| Subsequent LFDF | <11 | 80 (7.0%) | -- |
| Subsequent BMAC/PRP | <11 | <11 | -- |
| Specialized care for wound healing (inpatient or outpatient) | <11 | 24 (2.1%) | -- |
| Post-procedural complications inpatient care | <11 | 44 (3.8%) | -- |
| Cerebrospinal fluid leak inpatient care | <11 | <11 | -- |
| Mechanical device complications inpatient care | <11 | <11 | -- |
| Hematoma inpatient care | <11 | <11 | -- |
| Wound infection inpatient care | <11 | 16 (1.4%) | -- |
| Sepsis inpatient care | <11 | 16 (1.4%) | -- |
| Deep vein thrombosis inpatient care | <11 | <11 | -- |
| Pulmonary embolism inpatient care | <11 | 15 (1.3%) | -- |
| Other post-surgical embolism inpatient care | <11 | <11 | -- |
| DVT, PT, or OE inpatient care | <11 | 16 (1.4%) | -- |
| SNF services | <11 | <11 | -- |
| IRF services | <11 | <11 | -- |
| Home health services | <11 | 575 (50.0%) | < 0.001 |
| Opioid medication | 18 (43.9%) | 549 (47.7%) | 0.75 |

SD, standard deviation; MRI, magnetic resonance imaging; CT, computed tomography; LFDF, laminectomy, foraminotomy, discectomy, and facetectomy; BMAC, bone marrow aspirate concentrate; PRP, platelet rich plasma; CSF, cerebrospinal fluid; VDT, deep vein thrombosis; PE, pulmonary embolism; OE, obstructive event; SNF, skilled nursing facility; IRF, inpatient rehabilitation facility; Rx, prescription.

**Supplementary Table 5. Healthcare resource utilization 48 months post procedure – PRP versus fusion**

| **Characteristic** | **PRP**<br>N = 22 | **Fusion**<br>N = 724 | **p-value** |
|---|---|---|---|
| Number of outpatient visits | | | 0.9 |
| Mean (SD) | 14.5 (14.0) | 13.8 (14.5) | |
| Median (Q1, Q3) | 9.0 (4.0, 22.0) | 9.0 (4.0, 19.0) | |
| Number of outpatient physical therapy visits | | | 0.96 |
| Mean (SD) | 10.3 (21.6) | 8.7 (15.8) | |
| Median (Q1, Q3) | 1.5 (0.0, 15.0) | 2.0 (0.0, 12.0) | |
| Spine x-ray | 11 (50.0%) | 696 (96.1%) | < 0.001 |
| MRI of spine | 11 (50.0%) | 323 (44.6%) | 0.78 |
| CT of spine | <11 | 202 (27.9%) | 0.09 |
| Subsequent fusion | <11 | 98 (13.5%) | -- |
| Subsequent LFDF | <11 | 57 (7.9%) | -- |
| Subsequent BMAC/PRP | <11 | <11 | -- |
| Specialized care for wound healing (inpatient or outpatient) | <11 | 22 (3.0%) | -- |
| Post-procedural complications inpatient care | <11 | 29 (4.0%) | -- |
| Cerebrospinal fluid leak inpatient care | <11 | <11 | -- |
| Mechanical device complications inpatient care | <11 | <11 | -- |
| Hematoma inpatient care | <11 | <11 | -- |
| Wound infection inpatient care | <11 | 13 (1.8%) | -- |
| Sepsis inpatient care | <11 | 17 (2.3%) | -- |
| Deep vein thrombosis inpatient care | <11 | <11 | -- |
| Pulmonary embolism inpatient care | <11 | 12 (1.7%) | -- |
| Other post-surgical embolism inpatient care | <11 | <11 | -- |
| DVT, PT, or OE inpatient care | <11 | 14 (1.9%) | -- |
| SNF services | <11 | <11 | -- |
| IRF services | <11 | <11 | -- |
| Home health services | <11 | 375 (51.8%) | 0.04 |
| Opioid medication | 11 (50.0%) | 366 (50.6%) | > 0.99 |

SD, standard deviation; MRI, magnetic resonance imaging; CT, computed tomography; LFDF, laminectomy, foraminotomy, discectomy, and facetectomy; BMAC, bone marrow aspirate concentrate; PRP, platelet rich plasma; CSF, cerebrospinal fluid; VDT, deep vein thrombosis; PE, pulmonary embolism; OE, obstructive event; SNF, skilled nursing facility; IRF, inpatient rehabilitation facility; Rx, prescription.

**Supplementary Table 6. Costs – PRP versus fusion at 36- and 48-Months Post-Procedure**

| Characteristic | PRP<br>N = 41 | Fusion<br>N = 1,150 | Estimated mean difference (95% CI) | p-value |
|---|---|---|---|---|
| **36 months post-procedure** | | | | |
| External costs | | | | |
| Mean (SD) | 12,874 (10,143) | 43,347 (19,149) | 30,473 (27,137, 33,809) | <0.001 |
| Median (Q1, Q3) | 8,787 (8,162, 14,187) | 36,430 (35,450, 38,790) | | |
| External inflated costs | | | | |
| Mean (SD) | 16,994 (19,747) | 86,694 (38,299) | 69,700 (63,185, 76,214) | <0.001 |
| Median (Q1, Q3) | 9,937 (8,687, 17,834) | 72,860 (70,900, 77,580) | | |
| Commercial payer costs | | | | |
| Mean (SD) | 18,506 (17,978) | 67,628 (35,737) | 49,122 (43,174, 55,070) | <0.001 |
| Median (Q1, Q3) | 11,501 (9,397, 20,058) | 54,378 (51,012, 62,946) | | |
| Aggregate costing method | | | | |
| Mean (SD) | 28,404 (45,293) | 76,805 (58,059) | 48,401 (33,954, 62,848) | <0.001 |
| Median (Q1, Q3) | 13,121 (9,338, 29,779) | 63,615 (43,540, 94,837) | | |
| Aggregate costing method - truncated | | | | |
| Mean (SD) | 26,501 (29,091) | 72,852 (41,783) | 46,351 (37,008, 55,694) | <0.001 |
| Median (Q1, Q3) | 13,121 (11,443, 29,779) | 63,615 (43,540, 94,837) | | |
| **48 months post-procedure** | | | | |
| | **PRP**<br>N = 22 | **Fusion**<br>N = 724 | | |
| External costs | | | | |
| Mean (SD) | 14,193 (12,599) | 45,040 (21,597) | 30,847 (25,224, 36,471) | <0.001 |
| Median (Q1, Q3) | 9,582 (8,187, 14,217) | 36,695 (35,545, 39,795) | | |
| External inflated costs | | | | |
| Mean (SD) | 19,360 (24,119) | 90,080 (43,194) | 70,720 (59,917, 81,523) | <0.001 |
| Median (Q1, Q3) | 11,527 (8,737, 20,797) | 73,390 (71,090, 79,590) | | |
| Commercial payer costs | | | | |
| Mean (SD) | 21,467 (20,093) | 71,100 (40,167) | 49,634 (40,540, 58,727) | <0.001 |
| Median (Q1, Q3) | 14,107 (9,960, 28,039) | 54,893 (51,132, 68,551) | | |
| Aggregate costing method | | | | |
| Mean (SD) | 23,172 (15,618) | 82,053 (66,203) | 58,881 (50,627, 67,135) | <0.001 |
| Median (Q1, Q3) | 17,695 (9,935, 34,913) | 65,679 (45,529, 99,192) | | |
| Aggregate costing method - truncated | | | | |
| Mean (SD) | 24,863 (14,164) | 77,060 (44,904) | 52,197 (45,299, 59,094) | <0.001 |
| Median (Q1, Q3) | 17,695 (14,165, 34,913) | 65,679 (45,529, 99,192) | | |

**Supplementary Table 7. Healthcare Resource Utilization 36 months post procedure – PRP versus LFDF**

| Characteristic | PRP<br>N = 77 | LFDF<br>N = 1,953 | p-value |
|---|---|---|---|
| Number of outpatient visits | | | 0.01 |
| Mean (SD) | 9.6 (11.1) | 7.3 (9.7) | |
| Median (Q1, Q3) | 6.0 (3.0, 15.0) | 4.0 (1.0, 10.0) | |
| Number of outpatient physical therapy visits | | | 0.27 |
| Mean (SD) | 11.4 (23.5) | 7.4 (15.9) | |
| Median (Q1, Q3) | 2.0 (0.0, 12.0) | 1.0 (0.0, 9.0) | |
| Spine x-ray | 23 (29.9%) | 942 (48.2%) | 0.002 |
| MRI of spine | 24 (31.2%) | 756 (38.7%) | 0.22 |
| CT of spine | <11 | 164 (8.4%) | 0.7 |
| Subsequent fusion | <11 | 132 (6.8%) | 0.23 |
| Subsequent LFDF | <11 | 492 (25.2%) | < 0.001 |
| Subsequent BMAC/PRP | 11 (14.3%) | <11 | -- |
| Specialized care for wound healing (inpatient or outpatient) | <11 | 38 (1.9%) | -- |
| Post-procedural complications inpatient care | <11 | 41 (2.1%) | -- |
| Cerebrospinal fluid leak inpatient care | <11 | 12 (0.6%) | -- |
| Mechanical device complications inpatient care | <11 | <11 | -- |
| Hematoma inpatient care | <11 | <11 | -- |
| Wound infection inpatient care | <11 | 28 (1.4%) | -- |
| Sepsis inpatient care | <11 | 17 (0.9%) | -- |
| Deep vein thrombosis inpatient care | <11 | 12 (0.6%) | -- |
| Pulmonary embolism inpatient care | <11 | 11 (0.6%) | -- |
| Other post-surgical embolism inpatient care | <11 | <11 | -- |
| DVT, PT, or OE inpatient care | <11 | 16 (0.8%) | -- |
| SNF services | <11 | <11 | -- |
| IRF services | <11 | <11 | -- |
| Home health services | 17 (22.1%) | 605 (31.0%) | 0.12 |
| Opioid medication | 34 (44.2%) | 742 (38.0%) | 0.33 |

SD, standard deviation; MRI, magnetic resonance imaging; CT, computed tomography; LFDF, laminectomy, foraminotomy, discectomy, and facetectomy; BMAC, bone marrow aspirate concentrate; PRP, platelet rich plasma; CSF, cerebrospinal fluid; VDT, deep vein thrombosis; PE, pulmonary embolism; OE, obstructive event; SNF, skilled nursing facility; IRF, inpatient rehabilitation facility; Rx, prescription.

**Supplementary Table 8. Healthcare Resource Utilization 48 months post procedure – PRP versus LFDF**

| Characteristic | PRP<br>N = 50 | LFDF<br>N = 1,271 | p-value |
|---|---|---|---|
| Number of outpatient visits | | | 0.02 |
| Mean (SD) | 12.4 (13.5) | 9.1 (12.8) | |
| Median (Q1, Q3) | 6.5 (3.0, 19.0) | 4.0 (1.0, 12.0) | |
| Number of outpatient physical therapy visits | | | 0.74 |
| Mean (SD) | 11.4 (22.9) | 8.7 (19.1) | |
| Median (Q1, Q3) | 2.0 (0.0, 12.0) | 1.0 (0.0, 10.0) | |
| Spine x-ray | 17 (34.0%) | 662 (52.1%) | 0.02 |
| MRI of spine | 17 (34.0%) | 545 (42.9%) | 0.27 |
| CT of spine | <11 | 122 (9.6%) | -- |
| Subsequent fusion | <11 | 100 (7.9%) | -- |
| Subsequent LFDF | <11 | 333 (26.2%) | 0.002 |
| Subsequent BMAC/PRP | <11 | <11 | -- |
| Specialized care for wound healing (inpatient or outpatient) | <11 | 30 (2.4%) | -- |
| Post-procedural complications inpatient care | <11 | 27 (2.1%) | -- |
| Cerebrospinal fluid leak inpatient care | <11 | 11 (0.9%) | -- |
| Mechanical device complications inpatient care | <11 | <11 | -- |
| Hematoma inpatient care | <11 | <11 | -- |
| Wound infection inpatient care | <11 | 20 (1.6%) | -- |
| Sepsis inpatient care | <11 | 14 (1.1%) | -- |
| Deep vein thrombosis inpatient care | <11 | <11 | -- |
| Pulmonary embolism inpatient care | <11 | 12 (0.9%) | -- |
| Other post-surgical embolism inpatient care | <11 | <11 | -- |
| DVT, PT, or OE inpatient care | <11 | 16 (1.3%) | -- |
| SNF services | <11 | <11 | -- |
| IRF services | <11 | <11 | -- |
| Home health services | 16 (32.0%) | 439 (34.5%) | 0.83 |
| Opioid medication | 26 (52.0%) | 510 (40.1%) | 0.13 |

SD, standard deviation; MRI, magnetic resonance imaging; CT, computed tomography; LFDF, laminectomy, foraminotomy, discectomy, and facetectomy; BMAC, bone marrow aspirate concentrate; PRP, platelet rich plasma; CSF, cerebrospinal fluid; VDT, deep vein thrombosis; PE, pulmonary embolism; OE, obstructive event; SNF, skilled nursing facility; IRF, inpatient rehabilitation facility; Rx, prescription.

**Supplementary Table 9. Costs – PRP versus LFDF at 36- and 48-Months Post-Procedure**

| Characteristic | PRP<br>N = 77 | LFDF<br>N = 1,953 | Estimated mean difference (95% CI) | p-value |
|---|---|---|---|---|
| **36 months post-procedure** | | | | |
| External costs | | | | |
| Mean (SD) | 12,927 (10,030) | 16,243 (15,006) | 3,315 (963, 5,668) | 0.006 |
| Median (Q1, Q3) | 9,277 (8,137, 13,417) | 10,080 (9,150, 18,100) | | |
| External inflated costs | | | | |
| Mean (SD) | 16,830 (19,170) | 32,486 (30,012) | 15,656 (11,143, 20,170) | <0.001 |
| Median (Q1, Q3) | 10,917 (8,637, 16,617) | 20,160 (18,300, 36,200) | | |
| Commercial payer costs | | | | |
| Mean (SD) | 17,946 (17,328) | 37,673 (28,775) | 19,727 (15,625, 23,829) | <0.001 |
| Median (Q1, Q3) | 12,990 (9,397, 19,181) | 24,954 (21,571, 43,134) | | |
| Aggregate costing method | | | | |
| Mean (SD) | 24,024 (39,566) | 26,938 (39,511) | 2,914 (-6,158, 11,986) | 0.529 |
| Median (Q1, Q3) | 12,400 (9,280, 18,153) | 16,523 (10,661, 27,927) | | |
| Aggregate costing method - truncated | | | | |
| Mean (SD) | 20,016 (18,421) | 23,985 (21,241) | 3,968 (-282, 8,218) | 0.067 |
| Median (Q1, Q3) | 12,400 (9,280, 18,153) | 16,523 (10,661, 27,927) | | |
| **48 months post-procedure** | | | | |
| | **PRP**<br>N = 50 | **LFDF**<br>N = 1,271 | | |
| External costs | | | | |
| Mean (SD) | 13,712 (11,517) | 17,452 (16,928) | 37,40 (381, 7,099) | 0.029 |
| Median (Q1, Q3) | 9,787 (8,187, 14,217) | 10,340 (9,200, 18,400) | | |
| External inflated costs | | | | |
| Mean (SD) | 18,413 (22,281) | 34,905 (33,856) | 16,491 (9,975, 23,008) | <0.001 |
| Median (Q1, Q3) | 11,937 (8,737, 16,274) | 20,680 (18,400, 36,800) | | |
| Commercial payer costs | | | | |
| Mean (SD) | 19,924 (19,462) | 40,308 (31,946) | 20,385 (14,654, 26,115) | <0.001 |
| Median (Q1, Q3) | 13,916 (9,960, 20,085) | 25,772 (21,912, 45,021) | | |
| Aggregate costing method | | | | |
| Mean (SD) | 21,657 (31,346) | 29,695 (46,729) | 8,038 (-1,115, 17,192) | 0.085 |
| Median (Q1, Q3) | 12,713 (9,365, 19,476) | 17,078 (10,814, 29,903) | | |
| Aggregate costing method - truncated | | | | |
| Mean (SD) | 19,164 (16,664) | 25,703 (23,768) | 6,539 (1,689, 11,389) | 0.008 |
| Median (Q1, Q3) | 12,713 (9,365, 19,476) | 17,078 (10,814, 29,903) | | |

**Supplementary Table 10. Sensitivity analysis cohort flowchart**

| Inclusion/exclusion criteria | PRP Procedures | Fusion Procedures | LFDF Procedures |
|---|---|---|---|
| Starting population: Receiving eligible procedures | 3470 | 184721 | 235989 |
| Procedure occurred between 1/1/2016 and 12/31/2023 | 3217 | 177371 | 228356 |
| Exclude patients who have multiple DOBs or sexes in the data | 3216 | 177368 | 228349 |
| Patients age 18 or older at the time of the procedure | 3205 | 176328 | 227538 |
| 12 months of continuous commercial health plan enrollment after the procedure | 1038 | 115902 | 160129 |
| At least one outpatient claim in the 12 months after the procedure | 997 | 115828 | 160039 |
| **Procedure is elective and preceded by a relevant clinical diagnosis** | | | |
| Filter out emergent procedures for complications arising in the hospital | 997 | 99250 | 153768 |
| **Exclude patients with clinical contra-indications** | | | |
| Inflammatory syndrome dx in the 12 months prior to the procedure | 796 | 72177 | 124227 |
| Ineligible connective tissue disorder dx in the 12 months prior to the procedure | 777 | 70805 | 122711 |
| Infectious arthropathy dx in the 12 months prior to the procedure | 777 | 70717 | 122629 |
| Traumatic injury not suitable for Regenexx dx in the 12 months prior to the procedure | 710 | 64596 | 114727 |
| Other spondylopathies dx in the 12 months prior to the procedure | 706 | 63004 | 113610 |
| Anterior spinal or vertebral artery compression syndrome dx in the 12 months prior to the procedure | 706 | 62869 | 113560 |
| Congenital, neuromuscular, or idiopathic spinal deformity dx in the 12 months prior to the procedure | 685 | 57900 | 110351 |
| Spine infection dx in the 12 months prior to the procedure | 685 | 57900 | 110351 |
| Cancer: malignant neoplasms related to bone, spine, or nervous system | 685 | 57531 | 110081 |
| **Exclude patients with prior treatment contra-indications** | | | |
| Exclude Regenexx patients who had corticosteroids in the prior 2 months | 626 | 57531 | 110081 |
| Previous spine surgery in the 12 months prior to the procedure | 622 | 52710 | 93297 |
| RFA of spine overall in the 12 months prior to the procedure | 612 | 51140 | 92014 |
| Intraoperative and post-procedural complications and disorders of the musculoskeletal system in the 12 months prior to the procedure | 596 | 43829 | 88683 |
| Received mab medication in the 3 months prior to the procedure | 593 | 43219 | 87819 |
| Exclude surgery patients who had Regenexx at any point | 593 | 43201 | 87763 |
| If a patient had multiple procedures, take the first one | 540 | 41927 | 86163 |

**Supplementary Table 11. PRPvs. Fusion vs. LFDF**

| Characteristic | PRP<br>N = 540 | Fusion<br>N = 41,927 | LFDF<br>N = 86,163 |
|---|---|---|---|
| Continuous commercial health plan enrollment after index procedure | | | |
| CE for 12 months | 540 (100.0%) | 41,927 (100.0%) | 86,163 (100.0%) |
| CE for 24 months | 341 (63.1%) | 27,857 (66.4%) | 60,603 (70.3%) |
| CE for 36 months | 228 (42.2%) | 18,772 (44.8%) | 42,753 (49.6%) |
| CE for 48 months | 145 (26.9%) | 12,482 (29.8%) | 29,612 (34.4%) |
| Sex | | | |
| Female | 237 (43.9%) | 21,701 (51.8%) | 34,236 (39.7%) |
| Male | 303 (56.1%) | 20,226 (48.2%) | 51,927 (60.3%) |
| Age | | | |
| Mean (SD) | 50.5 (10.9) | 53.9 (9.5) | 48.6 (11.9) |
| Median (Q1, Q3) | 52.0 (43.0, 59.0) | 56.0 (48.0, 61.0) | 50.0 (40.0, 58.0) |
| Geographic region | | | |
| Northeast | 107 (19.8%) | 7,091 (16.9%) | 15,361 (17.8%) |
| Midwest | 210 (38.9%) | 19,130 (45.6%) | 38,852 (45.1%) |
| South | 128 (23.7%) | 12,012 (28.6%) | 23,699 (27.5%) |
| West | 94 (17.4%) | 3,657 (8.7%) | 8,181 (9.5%) |
| Other | <11 | 37 (0.1%) | 70 (0.1%) |
| Conditions of the lumbar spine compatible with DDD within 12 months prior to the procedure | | | |
| Lumbar disc degeneration or spondylosis | 258 (47.8%) | 29,909 (71.3%) | 57,492 (66.7%) |
| Lumbar spondylolisthesis | 54 (10.0%) | 21,670 (51.7%) | 10,925 (12.7%) |
| Lumbar stenosis or foraminal narrowing | 116 (21.5%) | 28,841 (68.8%) | 52,734 (61.2%) |
| Lumbar myelopathy | <11 | 1,409 (3.4%) | 3,372 (3.9%) |
| Lumbar radiculopathy | 240 (44.4%) | 31,583 (75.3%) | 76,916 (89.3%) |
| Conditions of the lumbar spine not indicative of DDD within 12 months prior to the procedure | | | |
| Herniated disc/intervertebral disc displacement | 133 (24.6%) | 17,538 (41.8%) | 59,127 (68.6%) |
| Other lumbar/lumbosacral disc disorders | <11 | 1,044 (2.5%) | 2,438 (2.8%) |
| Low back pain | 253 (46.9%) | 28,110 (67.0%) | 61,624 (71.5%) |
| Conditions related to the spine (not specific to lumbar) within 12 months prior to the procedure | | | |
| Deforming dorsopathies | 14 (2.6%) | 5,847 (13.9%) | 4,599 (5.3%) |
| Kyphosis | <11 | 843 (2.0%) | 636 (0.7%) |
| Multi-section conditions compatible with DDD within 12 months prior to the procedure | | | |
| Disc degeneration | 170 (31.5%) | 22,311 (53.2%) | 40,921 (47.5%) |
| Disc degeneration - number of regions | | | |
| One region | 148 (27.4%) | 16,720 (39.9%) | 32,461 (37.7%) |
| Two regions | 22 (4.1%) | 5,375 (12.8%) | 8,193 (9.5%) |
| Three or more regions | <11 | 216 (0.5%) | 267 (0.3%) |
| None | 370 (68.5%) | 19,616 (46.8%) | 45,242 (52.5%) |

| | | | |
|---|---|---|---|
| Instability | 52 (9.6%) | 22,194 (52.9%) | 9,951 (11.5%) |
| Instability - number of regions | | | |
| One region | 41 (7.6%) | 15,415 (36.8%) | 8,514 (9.9%) |
| Two regions | <11 | 5,875 (14.0%) | 1,329 (1.5%) |
| Three or more regions | <11 | 904 (2.2%) | 108 (0.1%) |
| None | 488 (90.4%) | 19,733 (47.1%) | 76,212 (88.5%) |
| Spinal stenosis | 101 (18.7%) | 31,727 (75.7%) | 50,367 (58.5%) |
| Spinal stenosis - number of regions | | | |
| One region | 84 (15.6%) | 23,174 (55.3%) | 39,856 (46.3%) |
| Two regions | 15 (2.8%) | 7,388 (17.6%) | 9,424 (10.9%) |
| Three or more regions | <11 | 1,165 (2.8%) | 1,087 (1.3%) |
| None | 439 (81.3%) | 10,200 (24.3%) | 35,796 (41.5%) |
| Spondylosis | 192 (35.6%) | 27,243 (65.0%) | 42,734 (49.6%) |
| Spondylosis - number of regions | | | |
| One region | 128 (23.7%) | 18,231 (43.5%) | 31,369 (36.4%) |
| Two regions | 57 (10.6%) | 7,468 (17.8%) | 9,976 (11.6%) |
| Three or more regions | <11 | 1,544 (3.7%) | 1,389 (1.6%) |
| None | 348 (64.4%) | 14,684 (35.0%) | 43,429 (50.4%) |
| Radiculopathy | 228 (42.2%) | 33,663 (80.3%) | 75,250 (87.3%) |
| Radiculopathy - number of regions | | | |
| One region | 153 (28.3%) | 20,278 (48.4%) | 41,752 (48.5%) |
| Two regions | 60 (11.1%) | 11,075 (26.4%) | 28,703 (33.3%) |
| Three or more regions | 15 (2.8%) | 2,310 (5.5%) | 4,795 (5.6%) |
| None | 312 (57.8%) | 8,264 (19.7%) | 10,913 (12.7%) |
| Myelopathy | <11 | 4,045 (9.6%) | 4,472 (5.2%) |
| Myelopathy - number of regions | | | |
| One region | <11 | 3,861 (9.2%) | 4,345 (5.0%) |
| Two regions | <11 | 169 (0.4%) | 119 (0.1%) |
| Three or more regions | <11 | 15 (0.0%) | <11 |
| None | 532 (98.5%) | 37,882 (90.4%) | 81,691 (94.8%) |
| Musculoskeletal conditions (not exclusive to the spine) within 12 months prior to the procedure | | | |
| Poor bone quality | <11 | 999 (2.4%) | 797 (0.9%) |
| Osteoarthritis in other parts of the body | 102 (18.9%) | 8,836 (21.1%) | 11,836 (13.7%) |
| General comorbidity diagnoses within 12 months prior to the procedure | | | |
| Obesity | 59 (10.9%) | 11,339 (27.0%) | 19,143 (22.2%) |
| Diabetes | 32 (5.9%) | 7,404 (17.7%) | 10,970 (12.7%) |
| Depression/Anxiety/Bipolar | 98 (18.1%) | 11,251 (26.8%) | 18,277 (21.2%) |
| Medications within 12 months prior to the procedure | | | |
| Opioid or tramadol medication | 130 (24.1%) | 15,747 (37.6%) | 35,238 (40.9%) |
| NSAID medication | 107 (19.8%) | 13,085 (31.2%) | 28,999 (33.7%) |
| Neuropathic pain medication | 60 (11.1%) | 12,243 (29.2%) | 24,854 (28.8%) |
| Intervention history in the 12 months prior to the procedure | | | |
| Diagnostic or therapeutic injection (CPT 62322, 62323) | 16 (3.0%) | 5,843 (13.9%) | 13,076 (15.2%) |
| Injection or anesthetic or steroid (CPT 64483, 64484) | 41 (7.6%) | 9,850 (23.5%) | 25,381 (29.5%) |
| Oral corticosteroids | 93 (17.2%) | 13,110 (31.3%) | 35,428 (41.1%) |
| Physical therapy | 214 (39.6%) | 17,815 (42.5%) | 41,756 (48.5%) |
| Greater than 6 PT visits | 115 (21.3%) | 8,139 (19.4%) | 19,144 (22.2%) |

| | | | |
|---|---|---|---|
| Number of PT visits | | | |
| Mean (SD) | 4.4 (8.6) | 3.8 (7.9) | 4.4 (8.2) |
| Median (Q1, Q3) | 0.0 (0.0, 5.0) | 0.0 (0.0, 4.0) | 0.0 (0.0, 6.0) |
| Health seeking behavior in the 12 months prior to the procedure | | | |
| Number of outpatient visits | | | |
| Mean (SD) | 16.6 (15.2) | 21.7 (15.7) | 19.5 (14.8) |
| Median (Q1, Q3) | 12.5 (5.0, 24.0) | 18.0 (11.0, 28.0) | 16.0 (9.0, 26.0) |
| ER visit | 62 (11.5%) | 7,277 (17.4%) | 17,596 (20.4%) |
| Number of ER visits | | | |
| Mean (SD) | 0.2 (0.5) | 0.3 (0.8) | 0.3 (0.8) |
| Median (Q1, Q3) | 0.0 (0.0, 0.0) | 0.0 (0.0, 0.0) | 0.0 (0.0, 0.0) |
| Chiropractic visit | 119 (22.0%) | 6,367 (15.2%) | 20,585 (23.9%) |
| Number of chiropractic visit | | | |
| Mean (SD) | 2.2 (6.2) | 1.3 (4.8) | 2.1 (5.7) |
| Median (Q1, Q3) | 0.0 (0.0, 0.0) | 0.0 (0.0, 0.0) | 0.0 (0.0, 0.0) |
| Spine x-ray | 153 (28.3%) | 31,101 (74.2%) | 56,849 (66.0%) |
| Spine MRI | 226 (41.9%) | 34,387 (82.0%) | 74,753 (86.8%) |
| Spine x-ray or spine MRI | 263 (48.7%) | 37,514 (89.5%) | 78,676 (91.3%) |
| Other severity proxies in the 12 months prior to the procedure | | | |
| Spinal stenosis outpatient visit | 101 (18.7%) | 31,555 (75.3%) | 50,172 (58.2%) |
| Number of spinal stenosis outpatient visits | | | |
| Mean (SD) | 0.4 (1.3) | 2.6 (3.3) | 1.6 (2.5) |
| Median (Q1, Q3) | 0.0 (0.0, 0.0) | 2.0 (1.0, 4.0) | 1.0 (0.0, 2.0) |
| Disc degeneration outpatient visit | 170 (31.5%) | 22,228 (53.0%) | 40,803 (47.4%) |
| Number of disc degeneration outpatient visits | | | |
| Mean (SD) | 1.0 (3.0) | 1.6 (3.0) | 1.3 (2.8) |
| Median (Q1, Q3) | 0.0 (0.0, 1.0) | 1.0 (0.0, 2.0) | 0.0 (0.0, 1.0) |
| Instability outpatient visit | 52 (9.6%) | 22,103 (52.7%) | 9,912 (11.5%) |
| Number of instability outpatient visits | | | |
| Mean (SD) | 0.2 (1.2) | 1.8 (3.0) | 0.3 (1.2) |
| Median (Q1, Q3) | 0.0 (0.0, 0.0) | 1.0 (0.0, 3.0) | 0.0 (0.0, 0.0) |
| Osteoarthritis outpatient visit | 102 (18.9%) | 8,696 (20.7%) | 11,718 (13.6%) |
| Number of osteoarthritis outpatient visits | | | |
| Mean (SD) | 0.7 (2.5) | 0.6 (2.1) | 0.3 (1.5) |
| Median (Q1, Q3) | 0.0 (0.0, 0.0) | 0.0 (0.0, 0.0) | 0.0 (0.0, 0.0) |
| Less invasive intervention indicator (CPT 62322, 62323, 64483, 64484, GT 6 PT visits) | 153 (28.3%) | 18,662 (44.5%) | 43,920 (51.0%) |

CE, continuous enrollment; DDD, degenerative disc disease; SD, standard deviation; CPT, current procedural terminology; MRI, magnetic resonance imaging; PT, physical therapy

**Supplementary Table 12. Healthcare Resource Utilization Outcomes Sensitivity Analysis – 12 months**

| Characteristic | PRP<br>N = 540 | Fusion<br>N = 41,927 | p-value |
|---|---|---|---|
| Number of outpatient visits | | | < 0.001 |
| Mean (SD) | 3.0 (4.9) | 6.3 (5.7) | |
| Median (Q1, Q3) | 1.0 (0.0, 4.0) | 5.0 (3.0, 8.0) | |
| Number of outpatient physical therapy visits | | | < 0.001 |
| Mean (SD) | 4.0 (9.6) | 5.6 (10.3) | |
| Median (Q1, Q3) | 0.0 (0.0, 3.0) | 0.0 (0.0, 7.0) | |
| Spine x-ray | 73 (13.5%) | 39,627 (94.5%) | < 0.001 |
| MRI of spine | 80 (14.8%) | 9,933 (23.7%) | < 0.001 |
| CT of spine | 11 (2.0%) | 7,393 (17.6%) | < 0.001 |
| Subsequent fusion | <11 | 3,256 (7.8%) | < 0.001 |
| Subsequent LFDF | <11 | 1,532 (3.7%) | 0.01 |
| Subsequent BMAC/PRP | 54 (10.0%) | <11 | -- |
| Specialized care for wound healing (inpatient or outpatient) | <11 | 1,060 (2.5%) | < 0.001 |
| Post-procedural complications inpatient care | <11 | 940 (2.2%) | < 0.001 |
| Cerebrospinal fluid leak inpatient care | <11 | 162 (0.4%) | -- |
| Mechanical device complications inpatient care | <11 | 308 (0.7%) | -- |
| Hematoma inpatient care | <11 | 224 (0.5%) | -- |
| Wound infection inpatient care | <11 | 732 (1.7%) | 0.003 |
| Sepsis inpatient care | <11 | 492 (1.2%) | 0.13 |
| Deep vein thrombosis inpatient care | <11 | 323 (0.8%) | -- |
| Pulmonary embolism inpatient care | <11 | 294 (0.7%) | -- |
| Other post-surgical embolism inpatient care | <11 | 32 (0.1%) | -- |
| DVT, PT, or OE inpatient care | <11 | 507 (1.2%) | 0.11 |
| SNF services | <11 | 379 (0.9%) | -- |
| IRF services | <11 | 128 (0.3%) | -- |
| Home health services | 62 (11.5%) | 18,854 (45.0%) | < 0.001 |
| Opioid medication | 139 (25.7%) | 19,940 (47.6%) | < 0.001 |

SD, standard deviation; MRI, magnetic resonance imaging; CT, computed tomography; LFDF, laminectomy, foraminotomy, discectomy, and facetectomy; BMAC, bone marrow aspirate concentrate; PRP, platelet rich plasma; CSF, cerebrospinal fluid; VDT, deep vein thrombosis; PE, pulmonary embolism; OE, obstructive event; SNF, skilled nursing facility; IRF, inpatient rehabilitation facility; Rx, prescription.

**Supplementary Table 13. Healthcare Resource Utilization Outcomes Sensitivity Analysis – 24 months**

| Characteristic | PRP<br>N = 341 | Fusion<br>N = 27,857 | p-value |
|---|---|---|---|
| Number of outpatient visits | | | < 0.001 |
| Mean (SD) | 5.2 (7.7) | 9.8 (9.3) | |
| Median (Q1, Q3) | 2.0 (0.0, 6.0) | 7.0 (4.0, 13.0) | |
| Number of outpatient physical therapy visits | | | < 0.001 |
| Mean (SD) | 6.9 (15.9) | 7.0 (13.0) | |
| Median (Q1, Q3) | 0.0 (0.0, 6.0) | 1.0 (0.0, 9.0) | |
| Spine x-ray | 63 (18.5%) | 26,624 (95.6%) | < 0.001 |
| MRI of spine | 69 (20.2%) | 9,818 (35.2%) | < 0.001 |
| CT of spine | <11 | 6,604 (23.7%) | < 0.001 |
| Subsequent fusion | <11 | 2,951 (10.6%) | < 0.001 |
| Subsequent LFDF | 13 (3.8%) | 1,530 (5.5%) | 0.22 |
| Subsequent BMAC/PRP | 33 (9.7%) | <11 | -- |
| Specialized care for wound healing (inpatient or outpatient) | <11 | 902 (3.2%) | 0.001 |
| Post-procedural complications inpatient care | <11 | 1,006 (3.6%) | 0.002 |
| Cerebrospinal fluid leak inpatient care | <11 | 117 (0.4%) | -- |
| Mechanical device complications inpatient care | <11 | 280 (1.0%) | -- |
| Hematoma inpatient care | <11 | 165 (0.6%) | -- |
| Wound infection inpatient care | <11 | 552 (2.0%) | 0.02 |
| Sepsis inpatient care | <11 | 471 (1.7%) | 0.34 |
| Deep vein thrombosis inpatient care | <11 | 259 (0.9%) | -- |
| Pulmonary embolism inpatient care | <11 | 237 (0.9%) | -- |
| Other post-surgical embolism inpatient care | <11 | 23 (0.1%) | -- |
| DVT, PT, or OE inpatient care | <11 | 410 (1.5%) | -- |
| SNF services | <11 | 303 (1.1%) | -- |
| IRF services | <11 | 116 (0.4%) | -- |
| Home health services | 67 (19.6%) | 14,139 (50.8%) | < 0.001 |
| Opioid medication | 119 (34.9%) | 14,177 (50.9%) | < 0.001 |

SD, standard deviation; MRI, magnetic resonance imaging; CT, computed tomography; LFDF, laminectomy, foraminotomy, discectomy, and facetectomy; BMAC, bone marrow aspirate concentrate; PRP, platelet rich plasma; CSF, cerebrospinal fluid; VDT, deep vein thrombosis; PE, pulmonary embolism; OE, obstructive event; SNF, skilled nursing facility; IRF, inpatient rehabilitation facility; Rx, prescription.

**Supplementary Table 14. Healthcare Resource Utilization Outcomes Sensitivity Analysis – 36 months**

| Characteristic | PRP<br>N = 228 | Fusion<br>N = 18,772 | p-value |
|---|---|---|---|
| Number of outpatient visits | | | < 0.001 |
| Mean (SD) | 7.0 (10.7) | 12.8 (12.7) | |
| Median (Q1, Q3) | 3.0 (1.0, 8.0) | 9.0 (4.0, 17.0) | |
| Number of outpatient physical therapy visits | | | 0.07 |
| Mean (SD) | 8.2 (17.2) | 8.2 (15.6) | |
| Median (Q1, Q3) | 0.0 (0.0, 9.0) | 2.0 (0.0, 11.0) | |
| Spine x-ray | 59 (25.9%) | 18,054 (96.2%) | < 0.001 |
| MRI of spine | 53 (23.2%) | 7,993 (42.6%) | < 0.001 |
| CT of spine | <11 | 5,067 (27.0%) | < 0.001 |
| Subsequent fusion | <11 | 2,376 (12.7%) | < 0.001 |
| Subsequent LFDF | <11 | 1,318 (7.0%) | 0.16 |
| Subsequent BMAC/PRP | 23 (10.1%) | <11 | -- |
| Specialized care for wound healing (inpatient or outpatient) | <11 | 730 (3.9%) | 0.01 |
| Post-procedural complications inpatient care | <11 | 867 (4.6%) | 0.004 |
| Cerebrospinal fluid leak inpatient care | <11 | 97 (0.5%) | -- |
| Mechanical device complications inpatient care | <11 | 216 (1.2%) | -- |
| Hematoma inpatient care | <11 | 114 (0.6%) | -- |
| Wound infection inpatient care | <11 | 412 (2.2%) | -- |
| Sepsis inpatient care | <11 | 409 (2.2%) | -- |
| Deep vein thrombosis inpatient care | <11 | 214 (1.1%) | -- |
| Pulmonary embolism inpatient care | <11 | 179 (1.0%) | -- |
| Other post-surgical embolism inpatient care | <11 | 15 (0.1%) | -- |
| DVT, PT, or OE inpatient care | <11 | 328 (1.7%) | -- |
| SNF services | <11 | 225 (1.2%) | -- |
| IRF services | <11 | 85 (0.5%) | -- |
| Home health services | 62 (27.2%) | 10,364 (55.2%) | < 0.001 |
| Opioid medication | 89 (39.0%) | 10,014 (53.3%) | < 0.001 |

SD, standard deviation; MRI, magnetic resonance imaging; CT, computed tomography; LFDF, laminectomy, foraminotomy, discectomy, and facetectomy; BMAC, bone marrow aspirate concentrate; PRP, platelet rich plasma; CSF, cerebrospinal fluid; VDT, deep vein thrombosis; PE, pulmonary embolism; OE, obstructive event; SNF, skilled nursing facility; IRF, inpatient rehabilitation facility; Rx, prescription.

**Supplementary Table 15. Healthcare Resource Utilization Outcomes Sensitivity Analysis – 48 months**

| **Characteristic** | **PRP**<br>N = 145 | **Fusion**<br>N = 12,482 | **p-value** |
|---|---|---|---|
| Number of outpatient visits | | | < 0.001 |
| Mean (SD) | 9.4 (13.9) | 15.8 (16.0) | |
| Median (Q1, Q3) | 4.0 (1.0, 11.0) | 10.0 (5.0, 21.0) | |
| Number of outpatient physical therapy visits | | | 0.17 |
| Mean (SD) | 9.1 (18.0) | 9.3 (17.4) | |
| Median (Q1, Q3) | 1.0 (0.0, 12.0) | 2.0 (0.0, 12.0) | |
| Spine x-ray | 43 (29.7%) | 12,061 (96.6%) | < 0.001 |
| MRI of spine | 39 (26.9%) | 5,910 (47.3%) | < 0.001 |
| CT of spine | <11 | 3,697 (29.6%) | < 0.001 |
| Subsequent fusion | <11 | 1,789 (14.3%) | < 0.001 |
| Subsequent LFDF | <11 | 975 (7.8%) | 0.07 |
| Subsequent BMAC/PRP | 18 (12.4%) | <11 | -- |
| Specialized care for wound healing (inpatient or outpatient) | <11 | 580 (4.6%) | 0.04 |
| Post-procedural complications inpatient care | <11 | 685 (5.5%) | 0.02 |
| Cerebrospinal fluid leak inpatient care | <11 | 76 (0.6%) | -- |
| Mechanical device complications inpatient care | <11 | 155 (1.2%) | -- |
| Hematoma inpatient care | <11 | 81 (0.6%) | -- |
| Wound infection inpatient care | <11 | 314 (2.5%) | -- |
| Sepsis inpatient care | <11 | 357 (2.9%) | -- |
| Deep vein thrombosis inpatient care | <11 | 190 (1.5%) | -- |
| Pulmonary embolism inpatient care | <11 | 157 (1.3%) | -- |
| Other post-surgical embolism inpatient care | <11 | 12 (0.1%) | -- |
| DVT, PT, or OE inpatient care | <11 | 289 (2.3%) | -- |
| SNF services | <11 | 168 (1.3%) | -- |
| IRF services | <11 | 74 (0.6%) | -- |
| Home health services | 48 (33.1%) | 7,318 (58.6%) | < 0.001 |
| Opioid medication | 66 (45.5%) | 7,022 (56.3%) | 0.01 |

SD, standard deviation; MRI, magnetic resonance imaging; CT, computed tomography; LFDF, laminectomy, foraminotomy, discectomy, and facetectomy; BMAC, bone marrow aspirate concentrate; PRP, platelet rich plasma; CSF, cerebrospinal fluid; VDT, deep vein thrombosis; PE, pulmonary embolism; OE, obstructive event; SNF, skilled nursing facility; IRF, inpatient rehabilitation facility; Rx, prescription.

**Supplementary Table 16. Healthcare Resource Utilization Outcomes Sensitivity Analysis – 12 months**

| **Characteristic** | **PRP**<br>N = 540 | **LFDF**<br>N = 86,163 | **p-value** |
|---|---|---|---|
| Number of outpatient visits | | | < 0.001 |
| Mean (SD) | 3.0 (4.9) | 3.7 (4.9) | |
| Median (Q1, Q3) | 1.0 (0.0, 4.0) | 2.0 (1.0, 5.0) | |
| Number of outpatient physical therapy visits | | | < 0.001 |
| Mean (SD) | 4.0 (9.6) | 4.5 (9.0) | |
| Median (Q1, Q3) | 0.0 (0.0, 3.0) | 0.0 (0.0, 5.0) | |
| Spine x-ray | 73 (13.5%) | 31,148 (36.2%) | < 0.001 |
| MRI of spine | 80 (14.8%) | 23,681 (27.5%) | < 0.001 |
| CT of spine | 11 (2.0%) | 4,109 (4.8%) | 0.004 |
| Subsequent fusion | <11 | 3,040 (3.5%) | < 0.001 |
| Subsequent LFDF | <11 | 17,378 (20.2%) | < 0.001 |
| Subsequent BMAC/PRP | 54 (10.0%) | <11 | -- |
| Specialized care for wound healing (inpatient or outpatient) | <11 | 1,298 (1.5%) | 0.007 |
| Post-procedural complications inpatient care | <11 | 1,101 (1.3%) | 0.01 |
| Cerebrospinal fluid leak inpatient care | <11 | 521 (0.6%) | -- |
| Mechanical device complications inpatient care | <11 | 28 (0.0%) | -- |
| Hematoma inpatient care | <11 | 310 (0.4%) | -- |
| Wound infection inpatient care | <11 | 1,059 (1.2%) | 0.02 |
| Sepsis inpatient care | <11 | 588 (0.7%) | -- |
| Deep vein thrombosis inpatient care | <11 | 356 (0.4%) | -- |
| Pulmonary embolism inpatient care | <11 | 293 (0.3%) | -- |
| Other post-surgical embolism inpatient care | <11 | 30 (0.0%) | -- |
| DVT, PT, or OE inpatient care | <11 | 528 (0.6%) | -- |
| SNF services | <11 | 256 (0.3%) | -- |
| IRF services | <11 | 134 (0.2%) | -- |
| Home health services | 62 (11.5%) | 18,477 (21.4%) | < 0.001 |
| Opioid medication | 139 (25.7%) | 34,795 (40.4%) | < 0.001 |

SD, standard deviation; MRI, magnetic resonance imaging; CT, computed tomography; LFDF, laminectomy, foraminotomy, discectomy, and facetectomy; BMAC, bone marrow aspirate concentrate; PRP, platelet rich plasma; CSF, cerebrospinal fluid; VDT, deep vein thrombosis; PE, pulmonary embolism; OE, obstructive event; SNF, skilled nursing facility; IRF, inpatient rehabilitation facility; Rx, prescription.

**Supplementary Table 17. Healthcare Resource Utilization Outcomes Sensitivity Analysis – 24 months**

| Characteristic | PRP<br>N = 341 | LFDF<br>N = 60,603 | p-value |
|---|---|---|---|
| Number of outpatient visits | | | < 0.001 |
| Mean (SD) | 5.2 (7.7) | 6.2 (8.3) | |
| Median (Q1, Q3) | 2.0 (0.0, 6.0) | 3.0 (1.0, 8.0) | |
| Number of outpatient physical therapy visits | | | 0.02 |
| Mean (SD) | 6.9 (15.9) | 5.9 (12.1) | |
| Median (Q1, Q3) | 0.0 (0.0, 6.0) | 0.0 (0.0, 7.0) | |
| Spine x-ray | 63 (18.5%) | 26,979 (44.5%) | < 0.001 |
| MRI of spine | 69 (20.2%) | 22,129 (36.5%) | < 0.001 |
| CT of spine | <11 | 4,551 (7.5%) | 0.002 |
| Subsequent fusion | <11 | 3,480 (5.7%) | 0.002 |
| Subsequent LFDF | 13 (3.8%) | 13,656 (22.5%) | < 0.001 |
| Subsequent BMAC/PRP | 33 (9.7%) | <11 | -- |
| Specialized care for wound healing (inpatient or outpatient) | <11 | 1,214 (2.0%) | 0.01 |
| Post-procedural complications inpatient care | <11 | 1,089 (1.8%) | 0.06 |
| Cerebrospinal fluid leak inpatient care | <11 | 397 (0.7%) | -- |
| Mechanical device complications inpatient care | <11 | 48 (0.1%) | -- |
| Hematoma inpatient care | <11 | 239 (0.4%) | -- |
| Wound infection inpatient care | <11 | 872 (1.4%) | -- |
| Sepsis inpatient care | <11 | 700 (1.2%) | -- |
| Deep vein thrombosis inpatient care | <11 | 344 (0.6%) | -- |
| Pulmonary embolism inpatient care | <11 | 285 (0.5%) | -- |
| Other post-surgical embolism inpatient care | <11 | 23 (0.0%) | -- |
| DVT, PT, or OE inpatient care | <11 | 517 (0.9%) | -- |
| SNF services | <11 | 242 (0.4%) | -- |
| IRF services | <11 | 107 (0.2%) | -- |
| Home health services | 67 (19.6%) | 17,120 (28.2%) | < 0.001 |
| Opioid medication | 119 (34.9%) | 27,372 (45.2%) | < 0.001 |

SD, standard deviation; MRI, magnetic resonance imaging; CT, computed tomography; LFDF, laminectomy, foraminotomy, discectomy, and facetectomy; BMAC, bone marrow aspirate concentrate; PRP, platelet rich plasma; CSF, cerebrospinal fluid; VDT, deep vein thrombosis; PE, pulmonary embolism; OE, obstructive event; SNF, skilled nursing facility; IRF, inpatient rehabilitation facility; Rx, prescription.

**Supplementary Table 18. Healthcare Resource Utilization Outcomes Sensitivity Analysis – 36 months**

| Characteristic | PRP<br>N = 228 | LFDF<br>N = 42,753 | p-value |
|---|---|---|---|
| Number of outpatient visits | | | 0.002 |
| Mean (SD) | 7.0 (10.7) | 8.3 (11.5) | |
| Median (Q1, Q3) | 3.0 (1.0, 8.0) | 4.0 (1.0, 11.0) | |
| Number of outpatient physical therapy visits | | | 0.71 |
| Mean (SD) | 8.2 (17.2) | 7.1 (14.8) | |
| Median (Q1, Q3) | 0.0 (0.0, 9.0) | 1.0 (0.0, 9.0) | |
| Spine x-ray | 59 (25.9%) | 21,462 (50.2%) | < 0.001 |
| MRI of spine | 53 (23.2%) | 18,063 (42.2%) | < 0.001 |
| CT of spine | <11 | 4,025 (9.4%) | 0.007 |
| Subsequent fusion | <11 | 3,076 (7.2%) | 0.01 |
| Subsequent LFDF | <11 | 10,326 (24.2%) | < 0.001 |
| Subsequent BMAC/PRP | 23 (10.1%) | <11 | -- |
| Specialized care for wound healing (inpatient or outpatient) | <11 | 1,061 (2.5%) | 0.08 |
| Post-procedural complications inpatient care | <11 | 942 (2.2%) | 0.11 |
| Cerebrospinal fluid leak inpatient care | <11 | 286 (0.7%) | -- |
| Mechanical device complications inpatient care | <11 | 61 (0.1%) | -- |
| Hematoma inpatient care | <11 | 167 (0.4%) | -- |
| Wound infection inpatient care | <11 | 710 (1.7%) | -- |
| Sepsis inpatient care | <11 | 653 (1.5%) | -- |
| Deep vein thrombosis inpatient care | <11 | 288 (0.7%) | -- |
| Pulmonary embolism inpatient care | <11 | 279 (0.7%) | -- |
| Other post-surgical embolism inpatient care | <11 | 22 (0.1%) | -- |
| DVT, PT, or OE inpatient care | <11 | 457 (1.1%) | -- |
| SNF services | <11 | 209 (0.5%) | -- |
| IRF services | <11 | 95 (0.2%) | -- |
| Home health services | 62 (27.2%) | 14,358 (33.6%) | 0.05 |
| Opioid medication | 89 (39.0%) | 20,850 (48.8%) | 0.004 |

SD, standard deviation; MRI, magnetic resonance imaging; CT, computed tomography; LFDF, laminectomy, foraminotomy, discectomy, and facetectomy; BMAC, bone marrow aspirate concentrate; PRP, platelet rich plasma; CSF, cerebrospinal fluid; VDT, deep vein thrombosis; PE, pulmonary embolism; OE, obstructive event; SNF, skilled nursing facility; IRF, inpatient rehabilitation facility; Rx, prescription.

**Supplementary Table 19. Healthcare Resource Utilization Outcomes Sensitivity Analysis – 48 months**

| **Characteristic** | **PRP**<br>N = 145 | **LFDF**<br>N = 29,612 | **p-value** |
|---|---|---|---|
| Number of outpatient visits | | | 0.06 |
| Mean (SD) | 9.4 (13.9) | 10.4 (14.7) | |
| Median (Q1, Q3) | 4.0 (1.0, 11.0) | 5.0 (2.0, 14.0) | |
| Number of outpatient physical therapy visits | | | 0.8 |
| Mean (SD) | 9.1 (18.0) | 8.2 (16.9) | |
| Median (Q1, Q3) | 1.0 (0.0, 12.0) | 1.0 (0.0, 10.0) | |
| Spine x-ray | 43 (29.7%) | 16,184 (54.7%) | < 0.001 |
| MRI of spine | 39 (26.9%) | 13,731 (46.4%) | < 0.001 |
| CT of spine | <11 | 3,229 (10.9%) | 0.01 |
| Subsequent fusion | <11 | 2,459 (8.3%) | 0.02 |
| Subsequent LFDF | <11 | 7,510 (25.4%) | < 0.001 |
| Subsequent BMAC/PRP | 18 (12.4%) | <11 | -- |
| Specialized care for wound healing (inpatient or outpatient) | <11 | 847 (2.9%) | --- |
| Post-procedural complications inpatient care | <11 | 766 (2.6%) | -- |
| Cerebrospinal fluid leak inpatient care | <11 | 230 (0.8%) | -- |
| Mechanical device complications inpatient care | <11 | 53 (0.2%) | -- |
| Hematoma inpatient care | <11 | 123 (0.4%) | -- |
| Wound infection inpatient care | <11 | 549 (1.9%) | -- |
| Sepsis inpatient care | <11 | 587 (2.0%) | -- |
| Deep vein thrombosis inpatient care | <11 | 261 (0.9%) | -- |
| Pulmonary embolism inpatient care | <11 | 249 (0.8%) | -- |
| Other post-surgical embolism inpatient care | <11 | 20 (0.1%) | -- |
| DVT, PT, or OE inpatient care | <11 | 415 (1.4%) | -- |
| SNF services | <11 | 175 (0.6%) | -- |
| IRF services | <11 | 84 (0.3%) | -- |
| Home health services | 48 (33.1%) | 11,237 (37.9%) | 0.27 |
| Opioid medication | 66 (45.5%) | 15,366 (51.9%) | 0.15 |

SD, standard deviation; MRI, magnetic resonance imaging; CT, computed tomography; LFDF, laminectomy, foraminotomy, discectomy, and facetectomy; BMAC, bone marrow aspirate concentrate; PRP, platelet rich plasma; CSF, cerebrospinal fluid; VDT, deep vein thrombosis; PE, pulmonary embolism; OE, obstructive event; SNF, skilled nursing facility; IRF, inpatient rehabilitation facility; Rx, prescription.
